# Disorders in lipid metabolism, oxidative stress, and antioxidants in patients with amnestic mild cognitive impairment without major depression

**DOI:** 10.1101/2024.06.08.24308614

**Authors:** Gallayaporn Nantachai, Michael Maes, Vinh-Long Tran-Chi, Arisara Amrapala, Asara Vasupanrajit, Solaphat Hemrungrojn, Chavit Tunvirachaisakul

## Abstract

**Background:** Amnestic mild cognitive impairment (aMCI) is characterized by changes in lipids and oxidative stress (OS). It is crucial to exclude patients with major depression (MDD) to accurately evaluate these biomarkers in aMCI.

**Aims:** To examine lipid and oxidative stress biomarkers associated with aMCI versus normal controls.

**Methods:** We performed a case-control analysis involving 61 individuals with aMCI (without MDD) and 60 healthy controls. We assessed the severity of aMCI, distress symptoms of old age, and lipid/OS biomarkers.

**Results:** The levels of serum -SH groups were significantly higher in individuals with aMCI, while the levels of malondialdehyde (MDA) were significantly lower in the same group. Serum advanced oxidation protein products, glutathione, and folic acid did not show any notable variations. In individuals with aMCI, we observed an elevated apolipoprotein B (ApoB)/ApoA ratio, as well as decreased levels of high-density lipoprotein cholesterol (HDL), ApoA, and a reverse cholesterol transport (RCT) index. The simultaneous presence of aMCI and subclinical depressive symptoms is marked by elevated levels of triglycerides and ApoB, as well as decreased levels of ApoA and HDL. A significant portion of the variability (24.9%) in a quantitative MCI severity score can be attributed to -SH groups, age (positively), MDA and education (inversely).

**Discussion:** The alterations in MDA and -SH levels in aMCI may potentially disrupt redox signaling, which can affect cell signaling and homeostatic setpoints. The interaction between aMCI and subclinical depressive symptoms can lead to increased atherogenicity and reduced antiatherogenic protection.

## 1. Introduction

Mild cognitive impairment (MCI) is the intermediate, transitional phase between dementia and typical aging, affecting 10-15% of people aged 65 and older (Anderson, 2019). MCI is identified by: (1) subjective cognitive complaints by the patient and/or informant; (2) poorer cognitive performance compared to normal adults of the same age and background as assessed via clinical objective evaluation; (3) reports of cognitive decline from the past year; (4) preserved general cognitive functioning with unaffected daily activities of living; and (5) absence of dementia (Morris, 2012; Portet et al., 2006). Currently, the global prevalence of MCI is around 19.7% (Song et al., 2023), and at least 10-15% of MCI cases progress to Alzheimer’s Dementia (AD) (Sanghuachang et al., 2023).

A meta-analysis reported the prevalence of major depressive disorder (MDD) in MCI is approximately 32% (Ismail et al., 2017), and 41 % in those with AD (Berk et al., 2023). Similarly, Kohler et al. (A Köhler et al., 2016) reported that 29.8% of people with MCI had depression, 18.3% had sleep disturbances, and 15.2% had apathy. A recent study (Tran-Chi et al., 2024) on MCI participants uncovered two separate dimensions within older adults who showed no signs of major depression (MDD). The first dimension is ‘mild cognitive dysfunctions,’ represented through a quantified MCI score (qMCI) indicating the extent of objective cognitive decline. The second dimension is referred to as ‘distress symptoms of old age (DSOA),’ which encompasses manifestations of anxiety, tension, and neuroticism. This dimension is linked to negative life events and adverse childhood experiences (ACEs).

Several studies report that blood biomarkers are associated with the pathology of MCI, including mechanisms involving lipids, oxidative stress, antioxidant defenses, and lipid peroxidation. Previous studies found changes in lipid profiles in aMCI and AD reminiscent of atherogenicity and lowered anti-atherogenic defenses, including increased triglycerides (TG), total cholesterol (TC), low-density lipoprotein cholesterol (LDL-C), and apolipoprotein B (ApoB), and lowered high-density lipoprotein cholesterol (HDL-C) and ApoA1 (Hong et al., 2022; Liu et al., 2020; Pillai et al., 2023; Tong et al., 2022; Weyman-Vela et al., 2022; Zuin et al., 2021). Additionally, an imbalance between lowered levels of detoxifying antioxidant defenses and increased reactive oxygen (ROS) and nitrogen (RNS) species has also been linked to MCI. For example, one recent meta-analysis reported increased oxidative stress toxicity due to increased lipid peroxidation, coupled with reduced levels of antioxidant defenses, including glutathione (GSH) in MCI compared to controls (Nantachai et al., 2022). In separate studies, malondialdehyde (MDA) accumulation that occurs with aging has been associated with an increased risk of both MCI and AD (Mangialasche et al., 2009), and plasma levels of advanced oxidation protein products (AOPP) were elevated in individuals with MCI and AD compared to controls (Chico et al., 2013). Furthermore, a simultaneous decline in plasma glutathione (GSH) levels and cognitive function was found (Lin and Lane, 2021). Lower serum total thiol (-SH) levels in MCI individuals were reported compared to controls (Gündüztepe et al., 2020). From another meta-analysis, AD patients had lower levels of folate and vitamins A, B12, C, and E compared to controls, indicating compromised antioxidant status (Pendlebury and Rothwell, 2009). However, the oxidative stress toxicity / antioxidant defenses (OSTOX/ANTIOX) and lipid correlates of the two dimensions of aMCI (qMCI and DSOA) have remained elusive.

Evidence has suggested that MDD patients have significant disorders in lipid metabolism and an increased OSTOX/ANTIOX ratio as well. A recent systematic review demonstrated increased atherogenicity biomarkers in MDD together with lowered lipid-associated antioxidant defenses and a reverse cholesterol transport (RCT) system with lowered lecithin-cholesterol acyl transferase (LCAT) activity and ApoA levels (Almulla et al., 2023). Additionally, MDD has been associated with low lecithin cholesterol acyl transferase (LCAT), high free cholesterol, and high ApoE levels compared to controls in another study (Maes et al., 2024b). Moreover, MDD patients were reported to have a higher Castelli risk index 1, which is the ratio between total cholesterol and high-density lipoprotein cholesterol (HDL-C) (Vargas et al., 2014). Importantly, previous reviews demonstrated that the oxidative and nitrosative stress (O&NS) processes are integral factors underlying MDD (Maes et al., 2011). Taken together, it can be deduced that the OSTOX/ANTIOX profiles of aMCI cannot be evaluated if patients with MDD are included in the same study sample. Subsequently, associations between the lipid and O&NS biomarkers, neurocognitive functioning, and qMCI and DSOA scores should be examined in aMCI after excluding MDD subjects to get a more accurate understanding of the relationship between these factors.

Hence, the objective of this study is to investigate the specific lipid and OSTOX/ANTIOX biomarkers of aMCI without MDD, and normal controls. Secondly, we will also examine the effects of these biomarkers on the severity of both aMCI (including qMCI scores) and DSOA scores. The specific hypotheses are that aMCI and DSOA are accompanied by (1) increased OSTOX (e.g., MDA and AOPP levels) and reduced ANTIOX (e.g. -SH groups, GSH, folic acid), and (2) increased atherogenicity (e.g., ApoB, Castelli risk index 1) but lowered antiatherogenic protection (e.g., HDL-C, ApoA, LCAT).

## 2. Materials and methods

### 2.1 Participants

An analysis was conducted to determine the required sample size, considering several factors such as the effect size, significance level, power, number of groups, and covariates. Based on this analysis, a sample size of 90 participants would be sufficient to conduct an ANCOVA with a power of 80%. To enhance the robustness of our analysis and account for potential dropouts, we decided to include an additional 30 subjects. As a result, the final study sample size consisted of 120 participants, all of whom were between the ages of 60 and 75.

Included in the study were 61 cases of aMCI referred from King Chulalongkorn Memorial Hospital in Thailand, along with 60 healthy controls. The two groups were selected from the same geographical region, specifically Bangkok, Thailand. Subjects with aMCI were recruited from various departments at King Chulalongkorn Memorial Hospital, including the Geriatric Clinic, the Cognitive Fitness Center unit, the Geriatric Psychiatry clinic, and the Neuroscience Center. The control group included a variety of individuals, such as senior Red Cross volunteers, attendees of Health Check-up Clinics, members of neighborhood senior organizations, and healthy elderly caregivers of aMCI patients who visited the Dementia Clinic.

An efficient screening process was conducted to identify eligible individuals for enrollment. Patients diagnosed with aMCI underwent assessment and met the criteria established by Petersen (2004). Additionally, they obtained a Clinical Dementia Rating (CDR) score of 0.5. These criteria encompass the identification of both subjective and objective memory impairments, the exclusion of dementia, and alterations in activities of daily living (ADL). Individuals with neurological conditions such as Parkinson’s disease, Alzheimer’s disease, multiple sclerosis, epilepsy, or stroke, were deemed ineligible for participation in the study, for both groups. In addition, the study excluded patients who had been diagnosed with various neuropsychiatric disorders such as substance use disorders, chronic fatigue syndrome, bipolar disorder, MDD, post-traumatic stress disorder, schizophrenia, obsessive-compulsive disorder, autism spectrum disorders, and generalized anxiety disorder, Ineligible for participation were individuals with medical conditions such as chronic kidney disease, cancer, metabolic syndrome, chronic inflammatory bowel disease, HIV infection, hepatitis, or chronic obstructive pulmonary disease. People with communication difficulties, visual impairments, hearing loss, physical conditions affecting their ability to sit or stand, or mobility issues were also not included.

The study received approval from the Institutional Review Board committee of the Faculty of Medicine Chulalongkorn University and the ethical committee of King Chulalongkorn Memorial Hospital (886/64). Prior to any data collection, all participants provided written informed consent.

### 2.2 Clinical measurements

This study utilized a case-control design, consisting of 60 control participants and 61 individuals with aMCI (Nantachai et al., 2023; Tran-Chi et al., 2024). We conducted a comprehensive interview to collect demographic and clinical information, utilizing a range of questionnaires. The Mini International Neuropsychiatric Interview (M.I.N.I.) (Kittirattanapaiboon, 2004) was utilized for the purpose of establishing axis-1 diagnoses, specifically MDD. To assess the extent of neurocognitive impairments, we employed the Montreal Cognitive Assessment (MoCA) scale (Hemrungrojn et al., 2021) and the Thai version of the Mini-Mental State Examination (MMSE) (Folstein et al., 1975). Additionally, we utilized the modified CDR score (Morris, 1993; Tran-Chi et al., 2024). The qMCI score, which serves as the primary principal component, was derived from the scores on the MoCA, MMSE, and the modified CDR score (Tran-Chi et al., 2024).

For the assessment of distress, depression, and anxiety symptoms, a variety of standardized scales were utilized. These included the Perceived Stress Scale (PSS) developed by Wongpakaran and Wongpakaran (2012), the State-Trait Anxiety Inventory (STAI) developed by Spielberger et al. (1971), the Thai Geriatric Depression Scale (TGDS) developed by Yesavage et al. (1982) and translate in Thai version by Pongwarin et.al, the depression (HADS-D) and anxiety (HADS-A) subscales of the Hospital Anxiety and Depression Scale developed by Nilchaikovit (1996), and the neuroticism trait score from the Five Factor Model standardized psychometric pool of items (IPIP-NEO) developed by Yomaboot and Cooper (2016). We organized the study group into smaller groups based on a HADS-D score of ≥8, resulting in two subgroups: one with higher depressive symptoms (n=26) and one with lower depressive symptoms (n=95). In a study conducted by Tran-Chi et al. (2024), a distress symptom of old age (DSOA) dimension was constructed through principal component analysis (PCA) on stress-affective symptoms. This dimension was derived as the first principal component extracted from various subscales including PSS, STAI, TGDS, HADS-D, HADS-A, and the neuroticism score. In previous research, it was found that there was no significant association between the DSOA and qMCI dimensional scores.

In addition, a specific group of individuals with cognitive impairments was identified through cluster analysis. This advanced machine learning technique effectively removed subjects who displayed symptoms associated with the DSOA dimension in aMCI subjects, as demonstrated in the study by Tran-Chi et al. (2024). Therefore, a more specific subset of aMCI, referred to as mCoDy (mild cognitive dysfunction), was established. In this study, we will discuss the findings from a group of individuals with aMCI (n = 61) as well as a more specific subgroup called mCoDy (n=52), which is more restricted and homogeneous. Furthermore, we employed the established Thai translation of the Adverse Childhood Experiences (ACEs) Questionnaire (Rungmueanporn et al., 2019)to evaluate five dimensions of ACE, including emotional, physical, and sexual abuse, as well as emotional and physical neglect. The Negative Event (Hassle) Scale, developed by Maybery et al. (2007), was utilized to assess negative life events (NLEs). The Alcohol, Smoking, and Substance Involvement Screening Test (ASSIST)(Humeniuk et al., 2008) was utilized to identify and eliminate individuals who engaged in substance use, including tobacco.

### 2.3 Assays

#### 2.3.1 Biological samples

Blood samples were obtained from the antecubital vein following an overnight fast between 8.00 and 9 a.m. Qualified nurses collected blood samples for the assay of lipids and O&NS biomarkers. All serum/plasma samples were kept at -80 °C until thawed for analyses. We employed the Alinity C (Abbott Laboratories, USA; Otawara-Shi, Tochigi-Ken, Japan) to assay total TC, HDL-C, TG, and direct LDL-C (Maes et al., 2024a). The inter-assay CV values were 2.3%, 2.6%, 2.3%, and 4.5%, for TC, HDL-C, TG, and LDL-C, respectively. ApoA1 and ApoB were measured using an immunoturbidimetric assay using the Roche Cobas 6000 and c501 module (Roche, Rotkreuz, Switzerland). The intra-assay CV values were 1.75% and 2.64% for ApoA1 and ApoB, respectively. Free cholesterol (FC) was assayed using the Free Cholesterol Colorimetric Assay Kit (Elabscience, cat number: E-BC-K004-M), as explained previously (Maes et al., 2024a). Based on these measurements, we determined the cholesterol esterification rate (CER) using the following formula: 1 – (free cholesterol / total cholesterol) x 100 (Maes et al. 2024a). Accordingly, the reverse cholesterol ratio was computed as: the z transformation of ApoA (zApoA) + z HDL-C + z CER (Maes et al., 2024a).

The Thiol Quantification Assay Kit, Fluorometric (Abcam, catalogue number: ab112158), was employed to assay free thiol. Briefly, 50 µl of the reaction mixture (1x thiol green indicator stock solution in the assay buffer) and 50 µl of the protein standards or samples were added to a solid black 96-well microplate. Then, the plate was incubated at room temperature for 60 minutes in the dark. The fluorescence increase was measured at Ex/Em = 490/520 nm with a microplate reader (Varioskan Flash Multimode, Thermo). A blank well was used as the control and was subtracted with the values of the protein standards and samples. The amount of free thiol in test samples was calculated from the values of the standard curve.

The MDA assay kit, Colorimetric (Abcam, catalogue number: ab118970), was used for measuring the lipid peroxidation in plasma. For sample preparation, 20 µl of plasma and 500 µl of 42 mM H2SO4 were added to a microcentrifuge tube. Then, 125 µL of phosphotungstic acid solution was added to their reaction, mixed using a vortex mixer, and incubated at room temperature for 5 minutes. The pellet was collected and suspended on ice with 200 µl of sterile water (with 2 µL BHT Stock/BHT (100X)). For the assay procedure, 200 µL of the developer VII/TBA reagent was added to each microcentrifuge tube containing 200 µL standard or 200 µL sample. The reaction mixes were heated at 95 °C for 60 minutes and then cooled to room temperature before the detection. The heated samples were taken into a clear 96-well plate with a flat bottom. The absorbance was measured at 532 nm using a microplate reader (Varioskan Flash Multimode, Thermo). The MDA concentration was calculated according to the manufacturer’s instructions and following the formula: 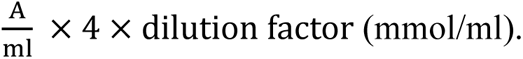

The presence of GSH in serum was detected using the GSH assay kit, Colorimetric (Abcam, catalogue number: ab239727). Briefly, 20 µl of diluted serums or standard proteins were added to a clear 96-well plate with a flat bottom. Then, 80 µl of the reaction mix was added to each well. The plate immediately measured the absorbance at 450 nm in kinetic mode at room temperature for 60 minutes using a microplate reader (Varioskan Flash Multimode, Thermo). For data analysis, the two time points (t1 and t2) in the linear range of the plot were selected and the corresponding absorbance values (OD1 and OD2) were obtained. The amount of GSH in test samples was calculated from the values of their standard curve following the calculation of the manufacturer’s instructions (Korzekwa et al., 2020; Tang et al., 2019).

The advanced oxidation protein products (AOPPs) in plasma were detected using the AOPP Assay Kit (Abcam, catalogue number: ab242295). 200 µl of test samples or standard proteins were prepared on a clear 96-well plate with a flat bottom. Then, 10 µl of Chloramine Reaction Initiator was added to each well and incubated on the orbital shaker at room temperature for 5 minutes. After incubation, the stop solution was added to each well. The absorbance was immediately read at 340 nm using a microplate reader (Varioskan Flash Multimode, Thermo). Data analysis was calculated from the standard curve. The standard curve was created for each assay performed.

### 2.4 Statistics

An analysis of variance was employed to investigate the variations in continuous variables among various diagnostic groups. Associations between the diagnosis of aMCI and the biomarkers were evaluated using multivariate and univariate GLM analysis. We implemented false discovery rate (FDR) p-correction on the biomarkers, which included multiple comparisons. In addition, we examined the effects of the interaction pattern between the diagnosis of aMCI X HADS-D groups. The chi-square test or Fisher’s Exact Probability Test was used to compare nominal variables across distinct categories. Stepwise automatic binary logistic regression analysis was performed with inclusion and exclusion p-values set at 0.05 and 0.06, respectively, to delineate the most relevant biomarkers (with demographic, ACE, and NES data) of the diagnosis aMCI (dependent variable with controls as reference group). We employed manual and automatic multiple regression analyses to determine the most accurate predictors of quantitative scores (e.g., qMCI, MMSE, DSOA) using biomarkers and demographic data. We also utilized a forward stepwise automatic regression method with inclusion and exclusion *p*-values set at 0.05 and 0.06, respectively. The final regression models included standardized coefficients, t-statistics, and exact *p*-values for each explanatory variable. Additionally, the total variance was represented by effect size measures such as R2 or partial eta squared, and F statistics (along with their corresponding *p*-values). We employed both the White and modified Breusch-Pagan test to evaluate for heteroskedasticity of the errors. An analysis was conducted to examine collinearity and multicollinearity. Various measures, such as tolerance (with a cut-off value of less than 0.25), the variance inflation factor (with a cut-off value of greater than 4), condition index, and variance proportions from the collinearity diagnostics table were utilized to assess the presence of heteroskedasticity. Furthermore, the data underwent z-score transformation to enhance their interpretability and we generated z-unit-weighted composite scores that accurately depict distinct immune profiles. The data distributions were subjected, where necessary, to various normalization processes, such as logarithmic or rank-based transformations. In each investigation mentioned, a two-tailed design was used to determine statistical significance at an α value of 0.05. The software used for analysis was IBM Windows SPSS version 29.

The a priori sample size was calculated using G*Power 3.1.9.4. The primary outcome analysis was a multiple regression analysis with the qMCI score as the dependent variable and biomarker data as the input variables. With a given f value of 0.179 (which corresponds to around 15% of the explained variance), 5 explanatory variables, an alpha value of 0.05, and a power of 0.8, the minimum sample size required was 78.

## 3. Results

### 3.1 Socio-demographic and clinical features of the MCI participants

**Table 1** displays the socio-demographic and clinical features of aMCI and control individuals in the current study. The MoCA and MMSE scores were significantly lower in aMCI subjects than in controls. The PSS score was slightly increased in aMCI, although there were no significant differences in the HADS depression and anxiety, STAI, and neuroticism scores among aMCI and control individuals. Emotional neglect and negative life events scores did not significantly differ between aMCI subjects and controls.

**Table. 1.** Clinical data of subjects with amnestic mild cognitive impairment (aMCI) and healthy controls (HC).

| Variables | HC (n=60) | aMCI (n=61) | F/X<sup>2</sup>/FET | df | P |
| --- | --- | --- | --- | --- | --- |
| Age (years) | 66.80 (4.07) | 68.72 (3.83) | 7.13 | 1/119 | 0.009 |
| Sex (male/female) | 13/47 | 17/44 | 0.62 | 1 | 0.430 <sup>a</sup> |
| Marital status (S/M/D) (n) | 15/36/9 | 14/40/7 | 0.48 | 2 | 0.784 <sup>a</sup> |
| Education (years) | 15.88 (2.94) | 13.70 (4.46) | 10.01 | 1/119 | 0.002 |
| Occupations (N/O/E/B/Ot) | 30/13/1/5/11 | 25/8/3/14/11 | 6.90 | 4 | 0.141<sup>a</sup> |
| BMI (kg/m<sup>2</sup>) | 22.66 (2.92) | 23.26 (3.50) | 1.07 | 1/119 | 0.303 |
| Total MoCA | 27.43 (1.51) | 22.33 (1.65) | 314.65 | 1/119 | <0.001 |
| Total MMSE | 28.70 (1.23) | 26.39 (2.27) | 47.73 | 1/119 | <0.001 |
| IPIP-NEO (neuroticism) | 14.33 (4.60) | 15.90 (5.13) | 3.12 | 1/119 | 0.080 |
| Perceived Stress scale | 12.25 (6.39) | 15.03 (5.59) | 6.49 | 1/119 | 0.012 |
| HADS-A | 4.53 (2.49) | 5.43 (3.12) | 3.00 | 1/119 | 0.085 |
| HADS-D | 3.97 (3.63) | 4.52 (3.77) | 0.68 | 1/118 | 0.411 |
| STAI | 36.10 (7.39) | 37.18 (8.81) | 0.53 | 1/119 | 0.467 |
| NLEs (health+money) | 0.84(0.13) | 0.88 (0.16) | 2.02 | 1/118 | 0.157 |
| NLEs (total) | 15.69 (14.72) | 14.91(12.94) | 0.09 | 1/118 | 0.759 |
| ACEs (neglect) | 18.00 (5.63) | 19.67(6.23) | 2.39 | 1/119 | 0.125 |
| 5ACEs (z score) | -0.19(0.89) | 0.19(1.06) | 4.86 | 1/119 | 0.029 |
| TGDS | 4.08 (3.16) | 6.18 (4.57) | 8.55 | 1/119 | 0.004 |
| ApoE (E2/E3, E3/E3, E3/E4) | 13/31/15 | 10/38/11 | 1.71 | 2/118 | 0.424 <sup>a</sup> |

All results are shown as means (SD). All results of analyses of variance (F), except <sup>a</sup>: $\chi^2$ – test, analysis of contingency analyses. Marital status; S: single, M: married, M: separate/divorced/widowed, Occupations (N/O/E/B/Ot) ; None, Government official, Employee, Private business, Other, MoCA: Montreal Cognitive Assessment, MMSE: Mini mental state examination, IPIP-NEO neuroticism: The International Personality Item Pool-NEO, neuroticism domain , HADS-A: Hospital Anxiety and Depression scale, anxiety scores, HADD-D: Hospital Anxiety and Depression scale, depression score, STAI: the State-Trait Anxiety Inventory, NLE: negative life events, ACEs (neglect): adverse childhood experiences (neglect), TGDS: Thai Geriatric Depression Scale, ApoE; apolipoprotein E (gene).

### 3.2 Differences between aMCI and controls

**Table 2** shows the results of the O&NS measurements. -SH groups were significantly higher in aMCI participants than in controls, whilst MDA was significantly lower in aMCI than in controls. There were no significant differences in folate, AOPP, and GSH (all results of GLM analysis with age, sex, BMI, and the drug status as covariates) and ApoE genotypes among the study groups No significant interactions between aMCI x HADS-D groups were found. The differences shown in **Table 2** remained significant after FDR p correction (at p<0.0125).

**Table. 2.** Lipids, oxidative stress, and antioxidant levels of subjects with amnestic mild cognitive impairment (aMCI) and healthy controls (HC).

| Variables | HC= 60 | aMCI=61 | F (F), X<sup>2</sup> | df | P |
| --- | --- | --- | --- | --- | --- |
| Folate (ng/ml) | 13.54 (0.70) | 13.87 (0.58) | 2.37* | 1/108 | 0.127 |
| Glutathione (reduced) (nmol/L) | 5.95 (0.65) | 4.87 (0.55) | 1.05* | 1/108 | 0.309 |
| -SH (thiol) groups (μmol/L) | 100.19 (6.26) | 133.20 (5.22) | 13.36* | 1/108 | **<0.001** |
| Malondialdehyde (μg/l) | 142.75 (8.62) | 123.38 (7.19) | 8.37* | 1/108 | **0.005** |
| Advanced Oxidation protein products (μmol/L) | 99.34 (5.68) | 94.17 (4.73) | 0.07* | 1/108 | 0.799 |
| ApoE (E2-E3 / E3-E3 / E3-E4) | 13/31/15 | 10/38/11 | 1.71 | 2 | 0.424<sup>a</sup> |
| Total cholesterol (mg/dl) | 209.98 (7.05) | 206.01 (5.88) | 0.39 (0.68) # | 1/104 | 0.535 (0.412) |
| Free cholesterol (mg/dl) | 40.37 (1.61) | 38.97 (1.34) | 0.23 (0.89) # | 1/104 | 0.636 (0.347) |
| Triglycerides (mg/dl) | 93.47 (7.90) | 99.41 (6.59) | 7.72 (0.04) # | 1/104 | **0.006** (0.844) |
| High density lipoprotein (mg/dl) | 64.07 (2.36) | 57.44 (1.97) | 7.63 (4.20) # | 1/104 | **0.007** (0.043) |
| Low density lipoprotein (mg/dl) | 124.14 (6.11) | 127.35 (5.09) | 2.81 (0.00) # | 1/104 | 0.097 (0.950) |
| Apolipoprotein A (μg/mL) | 166.80 (4.16) | 147.03 (3.47) | 6.26 (12.22) # | 1/104 | **0.014** (<0.001) |
| Apolipoprotein B (μg/mL) | 86.30 (3.88) | 91.08 (3.24) | 5.75 (0.54) # | 1/104 | **0.018** (0.466) |
| ApoB/ApoA ratio | 0.532 (0.033) | 0.636 (0.028) | 9.66 (6.90) # | 1/104 | **0.002** (0.010) |
| Lecithin-cholesterol acyltransferase (%) | 80.79 (0.51) | 80.99 (0.42) | 0.01 (0.11) # | 1/104 | 0.936 (0.742) |
| Reverse Cholesterol Transport index (z score) | 0.390 (0.171) | -0.144 (0.142) | 5.74 (5.24) # | 1/104 | **0.018** (0.024) |
| Castelli risk index 1 | 3.40 (3.71) | 3.71 (0.15) | 6.66 (1.38) # | 1/104 | **0.011** (0.243) |

All results are shown as means (SD). \*All results of GLM analyses after covarying for age, sex, body mass index and the drug status of the subjects. # All results of GLM analyses considering the interaction patterns between aMCI x HADS-D groups and after covarying for age, sex, body mass index and the drug status of the subjects. The first F value indicates the interaction pattern and the F value between brackets denotes the comparison between aMCI and controls. There were no significant differences in any of the biomarkers between the HADS-D groups. ApoE genotypes: results of $\chi^2$ – test, analysis of contingency analyses.
HADS-D: Hospital Anxiety and Depression scale, depression score.

We conducted a thorough investigation of the potential impact of the medicines and supplements that the participants were consuming on their oxidative stress profile. There were no significant differences in the use of antihypertensive drugs between the control group and the group with aMCI (43/17 versus 35/26, p=0.129). Similarly, there were no significant differences in the use of antidiabetic agents (54/6 versus 51/10, p=0.422), cholesterol lowering drugs (24/36 versus 25/36), vitamins (26/34 versus 31/30, p=0.409), fish oil (55/5 versus 51/10, p=0.270), and calcium supplements (40/20 versus 46/15, p=0.321). We have analyzed the potential impact of these medicines and supplements on the biomarkers of oxidative stress using multivariate and univariate GLM analysis, as presented in Table 1 of the **Electronic Supplementary File (ESF).** The multivariate GLM analysis revealed a significant impact of cholesterol-lowering drugs; the univariate GLM analysis indicated that this impact was specifically observed in the -SH groups, which were significantly reduced. This observed effect remained statistically significant after applying the FDR p correction at p=0.04.

The same table shows the measurements of TC, FC, triglycerides, LDL-C, ApoB, LCAT, and Castelli index in aMCI and control subjects. The results of GLM analyses, which were adjusted for age, sex, BMI, and drug status (as covariates), showed significant interaction patterns between aMCI and HADS-D groups. We found significant interaction patterns for triglycerides, HDL, ApoA, ApoB, ApoB/ApoA ratio, RCT, and CRI-I. All differences in the lipid markers shown in **Table 2** remained significant following FDR p correction (at *p*<0.0282). **ESF, Figures 1-7** illustrate the interaction patterns. While for people without depressive symptoms there were no significant differences in HDL, ApoA, and RCT between aMCI and control subjects, these three markers were lower in aMCI subjects with depressive symptoms compared to those without. While in people without depressive symptoms there were no significant differences in ApoB, ApoB/ApoA ratio, and Castelli risk index 1 between aMCI and control subjects, these markers were higher in subjects with higher HADS-D scores compared with their counterparts. HDL, ApoA, and the RCT index were significantly lower in aMCI than in controls, whilst the ApoB/ApoA ratio was significantly higher in aMCI than in controls.

We have analyzed the impact of the treatment administered to the participants on the lipid profile as presented in ESF, Table 2. The multivariate GLM analysis did not reveal any statistically significant effects on the lipid biomarkers. However, antidiabetic medications showed notable inhibitory effects on TC, LDL-C, ApoB, ApoB/ApoA ratio, and free cholesterol based on univariate GLM. The FDR p correction revealed that the impacts on TC, LDL-C, and ApoB remained statistically significant with a p-value of 0.0266. Regardless, we have calculated the residualized values for all biomarkers and assessed if utilizing these adjusted data will alter any outcomes. Nevertheless, there were no notable disparities observed when comparing the utilization of the unprocessed data with the residualized values.

**Table 3** shows the results of a binary logistic regression analysis when the aMCI diagnosis is the dependent variable with the healthy control group as reference group. Table 3, model 1 shows that aMCI was predicted by 4 variables, namely -SH groups (positively), education, MDA, and ApoB (all inversely) with a Nagelkerke value of 0.519 and an accuracy of 78.5%. We also examined whether the same variables would predict the restricted study group of mCoDy versus controls. Table 3, model 2 shows that mCoDy was predicted by the same variables with a Nagelkerke value of 0.456 and an accuracy of 79.2%. Model #3 shows that two variables predicted the subgroup with increased HADS-D scores, specifically AOPP and NES health+money with an accuracy of 79.5%.

**Table 3.** Results of binary logistic regression analyses with amnestic mild cognitive impairment (aMCI) or mild cognitive dysfunction (mCoDy) as the dependent variables and controls as reference group.

| Dependent variable | Explanatory Variables | B | SE | Wald | P | OR | 95% CI | X2 (df) | p | Nagelkerke |
| --- | --- | --- | --- | --- | --- | --- | --- | --- | --- | --- |
| **aMCI** | **Model #1** |  |  |  |  |  |  | 59.72 (4) | <0.001 | 0.519 |
|  | Education years | -0.185 | 0.071 | 6.83 | 0.009 | 0.83 | 0.72-0.96 |  |  |  |
|  | -SH groups | 1.279 | 0.298 | 18.40 | <0.001 | 3.59 | 2.00-6.43 |  |  |  |
|  | MDA | -2.728 | 0.740 | 13.60 | <0.001 | 0.07 | 0.02-0.28 |  |  |  |
|  | ApoA | -0.827 | 0.278 | 8.86 | 0.003 | 0.44 | 0.25-0.75 |  |  |  |
| **mCoDy** | **Model #2** |  |  |  |  |  |  | 44.36 (4) | <0.001 | 0.456 |
|  | Education years | -0.287 | 0.078 | 13.45 | <0.001 | 0.75 | 0.64-0.88 |  |  |  |
|  | -SH groups | 1.102 | 0.314 | 12.31 | <0.001 | 3.01 | 1.63-5.57 |  |  |  |
|  | MDA | -0.906 | 0.340 | 7.09 | 0.008 | 0.40 | 0.21-0.79 |  |  |  |
|  | ApoA | -0.858 | 0.296 | 8.40 | 0.004 | 0.42 | 0.24-0.76 |  |  |  |
| **High HADS-D** | **Model #3** |  |  |  |  |  |  | 14.46 (2) | 0.001 | 0.182 |
|  | AOPP | 0.529 | 0.238 | 4.93 | 0.026 | 1.70 | 1.06-2.71 |  |  |  |
|  | NLE_health+money | 0.780 | 0.258 | 9.13 | 0.003 | 2.18 | 1.32-3.62 |  |  |  |

-SH; Thiol groups, MDA; malondialdehyde, AOPP; advanced oxidation protein products, ApoA; apolipoprotein A,
NLE\_health+money: sum negative life events (health and money).

### 3.3 Results of multiple regression analysis

**Table 4** shows the results of multiple regression analyses with qMCI, MoCA, MMSE, DSAO, and HADS-D scores as dependent variables. Model #1 shows that 26.9% of the variance in the qMCI score was explained by the regression on education and MDA (both inversely) and -SH groups and age (both positively). Model #2 shows that 24.9% of the variance in the MoCA score was explained by the same four variables. We found that 13.9% of the variance in the MMSE score was explained by two variables, namely education and MDA (both positively). Finally, we also regressed the DSOA and HADS-D scores on all biomarkers, psychological data (ACE and NLE), and demographic data. Not one of the biomarker data was significantly associated with the DSOA or HADS-D scores. For example, model #4 shows that NLE health+money, education, and ACE neglect were associated with the DSAO score and explained 31.6% of its variance. Model #5 shows that NLE health+money, and ACE neglect were related to the HADS-D score and explained 16.4% of the variance.

**Table 4.**
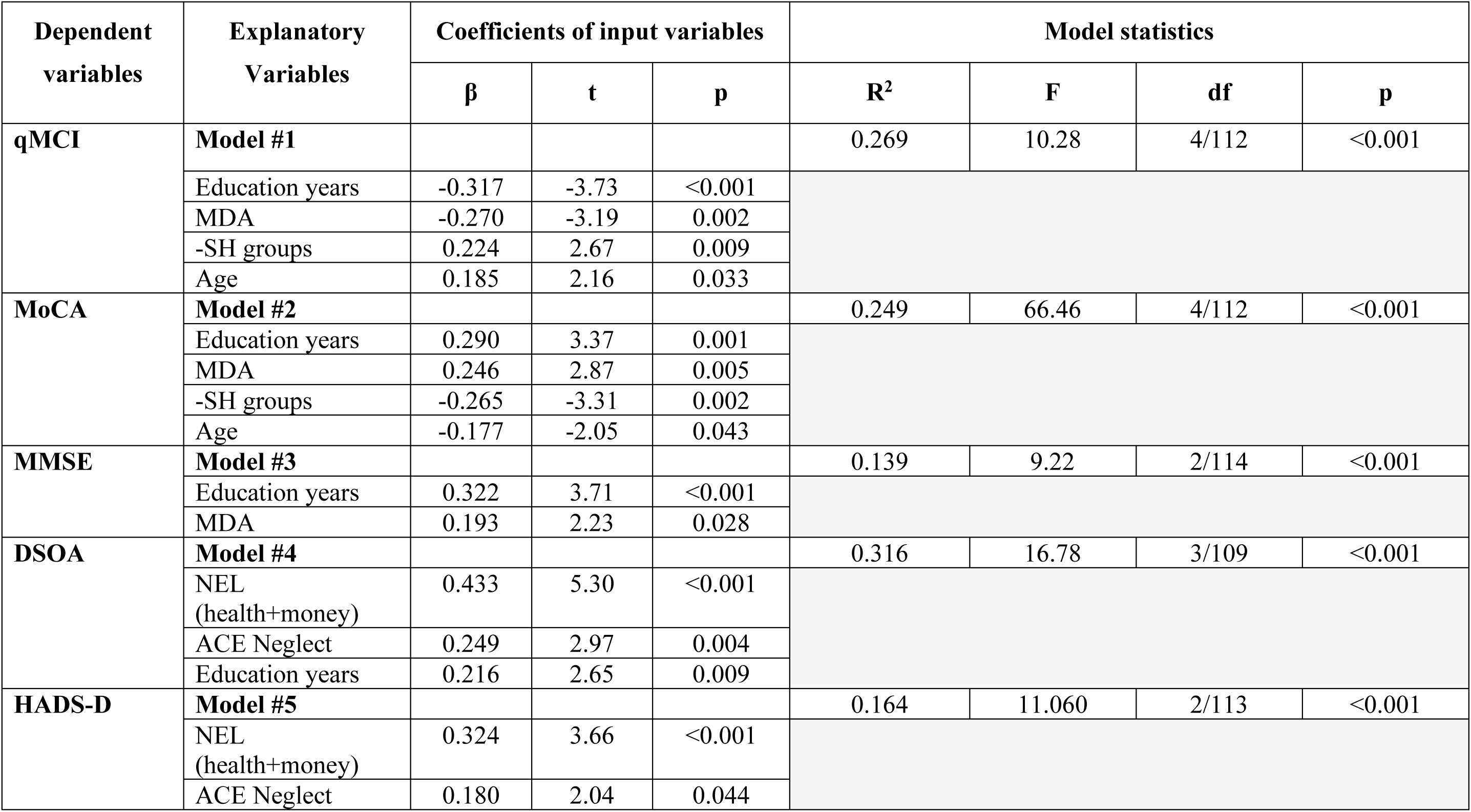
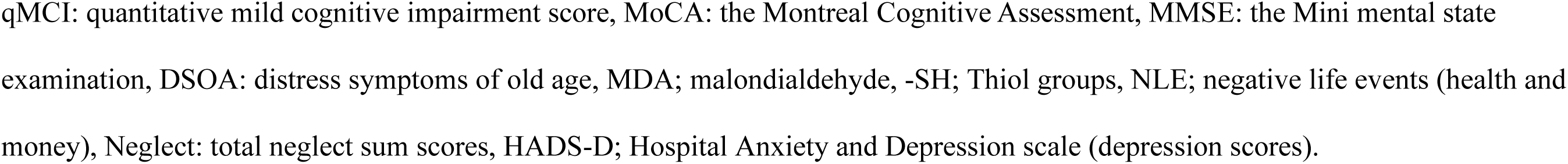
Results of multiple regression analysis with quantitative rating scores as dependent variables.

## 4. Discussion

### MDA in aMCI

Our study discovered that individuals with aMCI had lower MDA concentrations compared to the control group. Additionally, there were no notable variations in AOPP and antioxidants (GSH and folic acid) between the two groups. These findings challenge the initial hypothesis we had. According to a previous meta-analysis conducted by Nantachai et al. (2022), it was found that individuals with MCI had elevated levels of MDA and lipid hydroperoxides (LOOH) compared to the control group. The latter study examined elevated levels of other oxidative stress biomarkers, such as homocysteine and carbonyls, alongside reduced levels of antioxidant biomarkers, including vitamins, GSH, and GSH peroxidase (Nantachai et al., 2022).

One explanation for the differences is that subjects with MDD were not always excluded from the previous studies. MDD is known to be associated with higher levels of oxidative stress, such as increased MDA and AOPP levels, as well as lower levels of antioxidants like GSH and GSH peroxidase (Maes et al., 2011). Therefore, research on aMCI that includes patients with MDD may not provide a clear understanding of the true levels of oxidative stress and antioxidant biomarkers in aMCI.

MDA is formed through the degradation of arachidonic acid and other polyunsaturated fatty acids (Tsikas, 2017). It exhibits enhanced chemical stability and membrane permeability in comparison to reactive oxygen species (ROS) and demonstrates lower toxicity than other aldehyde compounds such as 4-hydroxy-2-nonenal (4-HNE) and methylglyoxal. In addition, several papers have proposed that MDA might offer protective effects, indicating that reduced MDA levels could potentially contribute to the pathophysiology of certain conditions. In fact, MDA can have a range of effects on the body, from potentially causing harm to offering protection.

MDA and 4-HNE function as signaling molecules that can stimulate gene expression and promote cell survival. However, they can also inhibit gene expression and contribute to cell death, as observed in studies conducted by Ayala et al. (2014). A recent study found that MDA acts as a signaling messenger, specifically influencing islet glucose-stimulated insulin secretion (GSIS) through the Wnt pathway. According to a study by Wang et al. (2014), it was discovered that MDA levels of 5 and 10 𝜇M have a moderate effect on enhancing islet GSIS. These levels were also found to increase the ATP/ADP ratio and cytosolic calcium levels, as well as influence gene expression and the production of proteins/activity associated with key regulators of GSIS.

The response of aldehyde dehydrogenase, which is activated by oxidative stress in mammals, is of utmost importance in determining the consequences of both immediate and prolonged elevations in aldehydes, such as MDA (Singh et al., 2014). The outcome is contingent upon several factors, including the extent and duration of MDA elevations, as well as the reaction of ALDH to oxidative stress. In a study conducted by Ayala et al. (2014), it was shown that lower levels of MDA could potentially interfere with important redox mechanisms in the body. Moreover, in the case of human beings, MDA represents a distinct epitope linked to oxidation, which is present on lipoproteins and cells that are undergoing apoptosis (Morris et al., 2019). This elicits an intrinsic immune response facilitated by IgM antibodies, which plays a crucial role in upholding homeostasis (Morris et al., 2019; Papac-Milicevic et al., 2016).

It is interesting to observe that within the plant kingdom, there is a captivating occurrence where short-lasting small increases in MDA levels can initiate a signaling process. This signaling process is of utmost importance in the regulation of cellular redox balance, the activation of the antioxidant system, and the plant’s ability to survive in challenging environments (Morales and Munné-Bosch, 2019). Continuous increases in MDA, on the other hand, lead to genotoxicity, a decrease in membrane fluidity, and various detrimental effects (Morales and Munné-Bosch, 2019). Based on this observation, it is possible that the decreased MDA concentrations in individuals with aMCI may be linked to a reduced ability of redox homeostatic processes.

### -SH groups and aMCI

Our study also unexpectedly revealed an elevation, rather than a reduction, in -SH groups among individuals diagnosed with aMCI. This association is consistent with a study that found higher thiol concentrations in MCI participants than in controls (Cervellati et al., 2014). Interestingly, elevated levels of thiol protein groups in individuals with depression are linked to reduced efficiency in declarative memory, working memory, and verbal fluency as well (Galecki et al., 2013).

None of our subjects took particular supplements known to elevate thiol groups (known as thiol supplements), such as lipoic acid, bromelain, glucosamine, selenium, cysteine, glutathione, and N-acetyl cysteine (Sen et al., 2000). Therefore, the increased levels of -SH groups in aMCI are not induced by intake of such supplements. The question is whether increased thiol accumulation may have a pathophysiological role and, if this were the case, why this could predispose individuals to develop aMCI. Enhanced -SH aggregation might coincide with heightened levels of S-prenylation (Morris et al., 2016) and increased glutathionylated protein levels (Musaogullari and Chai, 2020). Furthermore, an elevation in S-nitrosylation due to increased nitric oxide levels could also occur (Morris et al., 2016). The latter mechanisms may exert deleterious effects, potentially contributing to age-related processes and neurodegeneration, including AD (Musaogullari and Chai, 2020). It is important to emphasize that the prolonged administration of GSH or NAC to both young and aged animals may disrupt overall gene expression, suppress skn-1-mediated transcription, and hasten the aging process (Gusarov et al., 2021). On the contrary, restricting the intake of dietary thiols may prolong lifespan (Gusarov et al., 2021). Homocysteine, a thiol compound, constitutes a part of the overall plasma thiol pool, which comprises low molecular weight thiols, cysteine, glutathione, and cysteinylglycine (Garavaglia et al., 2022). Elevated levels of homocysteine are linked to neurocognitive deficits in older individuals and AD (Smith et al., 2018). Finally, high thiol antioxidant amounts could lead to increased oxidative damage, misfolded proteins, endoplasmic reticulum (ER) stress, and accelerated aging (Gusarov et al., 2021; Ravi et al., 2023).

### Lipid metabolism in aMCI

Our lipid profile results were more in line with the a priori hypothesis; there were lower HDL-C and ApoA concentrations, and a higher ApoB/ApoA ratio in aMCI participants compared to controls. Collectively, these findings indicate lowered antiatherogenic but increased atherogenesis processes. A prior meta-analysis examining correlations between lipid profiles and aMCI revealed that elevated TC and LDL-C levels were associated with increased cognitive impairment (Liu et al., 2020). There was a trend towards increased TC levels in aMCI, whilst there were no significant changes in LDL-C, TG, and HDL-C. Plasma TG levels were found to be higher in MCI than control subjects another case-control investigations (He et al., 2016; Tong et al., 2022; Weyman-Vela et al., 2022). Metabolic syndrome biomarkers (TG/HDL-C ratio and lowered ApoA1) correlated with a swifter deterioration of cognitive functions in subjects with aMCI (Pillai et al., 2023). Meta-analyses studies performed in AD showed that this illness is accompanied by increased TC and LDL-C (Liu et al., 2020), and lowered serum/plasma ApoA1(Tong et al., 2022; Zuin et al., 2021).

In addition to the observed associations between the lipid profiles and aMCI diagnosis, this study also found that the co-occurrence of aMCI and subclinical depressive symptoms was characterized by lowered HDL, ApoA, a reverse cholesterol transport index, and increased ApoB, ApoB/ApoA ratio, and a Castelli risk index 1. Increased ApoB/ApoA and Castelli risk index ratios are important predictors of increased risk of cardiovascular disease (Bhardwaj et al., 2013). Thus, subclinical depression already is sufficient in increasing atherogenicity when coupled with aMCI. Previous literature has mentioned that individuals diagnosed with MDD were found to exhibit significantly higher atherogenicity indices and a lowered reverse cholesterol transport (including HDL-C and ApoA1 levels) compared to control subjects (Bortolasci et al., 2015; Maes et al., 2024c; Nunes et al., 2015; Vargas et al., 2014). Moreover, Morelli et al. (Morelli et al., 2021) found that, in MDD, elevated atherosclerosis and insulin resistance were largely influenced by reactive oxygen and nitrogen species.

According to past research, atherogenicity is considered a risk factor for cognitive deficits and elevated atherogenic lipoproteins have been associated with impaired cognition (Reis et al., 1989). Additionally, low global cognitive performance, memory, and executive function have also been reported in those with atherogenic dyslipidemia (characterized by elevated triglyceride levels, increased LDL concentrations, and reduced HDL concentrations) (Ji et al., 2024). The relationship between dyslipidemia and cognitive deficits may be due to the role that dyslipidemia plays in the formation of atherosclerotic plaques, leading to ischemic injuries in the brain (Pendlebury and Rothwell, 2009).

## Limitations

This study was performed in Thai individuals with aMCI and, therefore, may not be generalizable to other ethnicities or countries. Therefore, the results deserved to be replicated in other countries and ethnicities.

## Conclusions

Collectively looking at our results, the study found decreased levels of MDA instead of increased MDA levels. In addition, there were no significant changes in antioxidant defenses, such as GSH, whilst the -SH groups were even increased. Although these results are contrary to our a priori hypothesis, it could potentially be explained by the underlying complex roles of MDA and -SH groups and different mechanisms of these biomarkers leading to alterations in homeostatic setpoints. Regarding the atherogenic lipid profile of aMCI, our study results agreed with the previous literature. Apart from this, we also report that the co-occurrence of aMCI with subclinical depressive symptoms could be characterized by increasing atherogenicity and lowered antiatherogenic protection. Studies that fail to exclude patients with MDD and neglect to control for subclinical depressive symptoms are unable to accurately determine the oxidative stress, antioxidant, and lipid profiles of aMCI.

## Supporting information

ESF, table 1-2, Figures 1-7

## Data Availability

All data produced in the present study are available upon reasonable request to the authors

## Acknowledgments

Not applicable.

## Disclosure

The authors declare no competing interests.

## Funding

The study was supported by the 90th Anniversary of Chulalongkorn University Scholarship under the Ratchadaphisek Somphot Fund Batch#52/2 (GCUGR1125652003D), and the Ratchadaphisek Somphot Fund (Faculty of Medicine), MDCU (Batch #GA65/34), The scholarship from graduate affair, Faculty of Medicine, Chulalongkorn University, Thailand, to GN. This research is funded by Thailand Science research and Innovation Fund Chulalongkorn University (HEA663000016) to MM.

## Credit authorship contribution statement

GN: data curation, investigation, project administration, first draft writing; MM: design, methodology, conceptualization, statistical analysis, visualization, writing, review, supervision; V-LT-C: data curation, investigation, project administration; AV: pre-data processing : AA, and SH: editing; CT: design, methodology, review, and editing. All authors approved the submitted draft of this paper.

## Availability of data

The corresponding author MM will provide the data file used in the present study upon receiving an appropriate request once the authors have fully utilized the data.

## Ethical approval and consent to participate

The research project (IRB no.886/64) received approval from the Institutional Review Board (IRB) at Chulalongkorn University’s ethics board in Bangkok, Thailand. This approval is in accordance with the International Guidelines for the Protection of Human Research Participants, as mandated by the Declaration of Helsinki, The Belmont Report, CIOMS Guidelines, and the International Conference on Harmonization in Good Clinical Practice (ICH-GCP). All participants signed the appropriate institutional informed consent forms before data collection. This study did not include any minors.

## Consent for publication

Not Applicable.

## Notes

### Funding Statement

This study was funded by by the 90th Anniversary of Chulalongkorn University Scholarship under the Ratchadaphisek Somphot Fund Batch#52/2 (GCUGR1125652003D), and the Ratchadaphisek Somphot Fund (Faculty of Medicine), MDCU (Batch #GA65/34), The scholarship from graduate affair, Faculty of Medicine, Chulalongkorn University, Thailand, to GN. This research is funded by Thailand Science research and Innovation Fund Chulalongkorn University (HEA663000016) to MM.

### Author Declarations

The Institutional Review Board of Chulalongkorn University ethics board, Thailand gave ethical approval for this research project.

