## Supplementary material for "Disorders in lipid metabolism, oxidative stress, and antioxidants in patients with amnestic mild cognitive impairment without major depression": ESF, table 1-2, Figures 1-7

### ELECTRONIC SUPPLEMENTARY FILE (ESF)

Running title: Lipid metabolism and oxidative stress in MCI

(1-3) Gallayaporn Nantachai*, (1,2,4-10) Michael Maes*, (2,10) Vinh-Long Tran-Chi, (2,6,10) Arisara Amrapala, (1,2) Asara Vasupanrajit, (2,6) Solaphat Hemrungrojn, (1,2,7,10) Chavit Tunvirachaisakul.

* Joint first authorships

1. Ph.D. program in Mental health, Department of Psychiatry, Faculty of Medicine, Chulalongkorn University, Bangkok, Thailand.

2. Department of Psychiatry, Faculty of Medicine, Chulalongkorn University, Bangkok, Thailand.

3. Somdet Phra Sungharaj Nyanasumvara Geriatric Hospital, Department of Medical Services, Ministry of Public Health, Chon Buri Province, Thailand.

4. Sichuan Provincial Center for Mental Health, Sichuan Provincial People’s Hospital, School of Medicine, University of Electronic Science and Technology of China, Chengdu 610072, China.

5. Key Laboratory of Psychosomatic Medicine, Chinese Academy of Medical Sciences, Chengdu, 610072, China.

6. Cognitive Fitness and Biopsychiatry Technology Research Unit, Faculty of Medicine, Chulalongkorn University, Bangkok, Thailand.

7. Center of Excellence in Cognitive Impairment and Dementia, Department of Psychiatry, Faculty of Medicine, Chulalongkorn University, Bangkok, Thailand.

8. Research Institute, Medical University of Plovdiv, Plovdiv, Bulgaria.

9. Department of Psychiatry, Medical University of Plovdiv, Plovdiv, Bulgaria.

10. Ph.D. Program in Clinical Sciences, School of Global Health, Faculty of Medicine, Chulalongkorn University, Bangkok, Thailand.

**ESF, Table 1.** Effects of medications and supplements on oxidative stress variables.

**Multivariate Tests[a]**

| Effect | | Value | F | Hypothesis df | Error df | Sig. | Partial Eta Squared |
|---|---|---|---|---|---|---|---|
| Intercept | Pillai's Trace | .165 | 4.028[b] | 5.000 | 102.000 | .002 | .165 |
| | Wilks' Lambda | .835 | 4.028[b] | 5.000 | 102.000 | .002 | .165 |
| | Hotelling's Trace | .197 | 4.028[b] | 5.000 | 102.000 | .002 | .165 |
| | Roy's Largest Root | .197 | 4.028[b] | 5.000 | 102.000 | .002 | .165 |
| Sex | Pillai's Trace | .126 | 2.939[b] | 5.000 | 102.000 | .016 | .126 |
| | Wilks' Lambda | .874 | 2.939[b] | 5.000 | 102.000 | .016 | .126 |
| | Hotelling's Trace | .144 | 2.939[b] | 5.000 | 102.000 | .016 | .126 |
| | Roy's Largest Root | .144 | 2.939[b] | 5.000 | 102.000 | .016 | .126 |
| Age | Pillai's Trace | .053 | 1.144[b] | 5.000 | 102.000 | .342 | .053 |
| | Wilks' Lambda | .947 | 1.144[b] | 5.000 | 102.000 | .342 | .053 |
| | Hotelling's Trace | .056 | 1.144[b] | 5.000 | 102.000 | .342 | .053 |
| | Roy's Largest Root | .056 | 1.144[b] | 5.000 | 102.000 | .342 | .053 |
| Lipid_drug | Pillai's Trace | .111 | 2.560[b] | 5.000 | 102.000 | .032 | .111 |
| | Wilks' Lambda | .889 | 2.560[b] | 5.000 | 102.000 | .032 | .111 |
| | Hotelling's Trace | .125 | 2.560[b] | 5.000 | 102.000 | .032 | .111 |
| | Roy's Largest Root | .125 | 2.560[b] | 5.000 | 102.000 | .032 | .111 |
| HT_drug | Pillai's Trace | .040 | .851[b] | 5.000 | 102.000 | .517 | .040 |
| | Wilks' Lambda | .960 | .851[b] | 5.000 | 102.000 | .517 | .040 |
| | Hotelling's Trace | .042 | .851[b] | 5.000 | 102.000 | .517 | .040 |
| | Roy's Largest Root | .042 | .851[b] | 5.000 | 102.000 | .517 | .040 |
| DM_drug | Pillai's Trace | .034 | .727[b] | 5.000 | 102.000 | .604 | .034 |
| | Wilks' Lambda | .966 | .727[b] | 5.000 | 102.000 | .604 | .034 |

| | | | | | | | |
|---|---|---|---|---|---|---|---|
| | Hotelling's Trace | .036 | .727[b] | 5.000 | 102.000 | .604 | .034 |
| | Roy's Largest Root | .036 | .727[b] | 5.000 | 102.000 | .604 | .034 |
| vitamins | Pillai's Trace | .047 | 1.016[b] | 5.000 | 102.000 | .412 | .047 |
| | Wilks' Lambda | .953 | 1.016[b] | 5.000 | 102.000 | .412 | .047 |
| | Hotelling's Trace | .050 | 1.016[b] | 5.000 | 102.000 | .412 | .047 |
| | Roy's Largest Root | .050 | 1.016[b] | 5.000 | 102.000 | .412 | .047 |
| Calcium | Pillai's Trace | .076 | 1.682[b] | 5.000 | 102.000 | .146 | .076 |
| | Wilks' Lambda | .924 | 1.682[b] | 5.000 | 102.000 | .146 | .076 |
| | Hotelling's Trace | .082 | 1.682[b] | 5.000 | 102.000 | .146 | .076 |
| | Roy's Largest Root | .082 | 1.682[b] | 5.000 | 102.000 | .146 | .076 |
| fish_oil | Pillai's Trace | .045 | .961[b] | 5.000 | 102.000 | .445 | .045 |
| | Wilks' Lambda | .955 | .961[b] | 5.000 | 102.000 | .445 | .045 |
| | Hotelling's Trace | .047 | .961[b] | 5.000 | 102.000 | .445 | .045 |
| | Roy's Largest Root | .047 | .961[b] | 5.000 | 102.000 | .445 | .045 |

Lipid drugs: cholesterol lowering drugs, HT drugs: antihypertensive drugs, DM drugs: antidiabetic agents.

**Tests of Between-Subjects Effects**

| Source | Dependent Variable | Type III Sum of Squares | df | Mean Square | F | Sig. | Partial Eta Squared |
|---|---|---|---|---|---|---|---|
| Corrected Model | Folate | 256.533[a] | 8 | 32.067 | 2.310 | .025 | .148 |
| | GSH | 56.364[b] | 8 | 7.045 | .568 | .802 | .041 |
| | SH_THIOL | 312.400[c] | 8 | 39.050 | 3.035 | .004 | .186 |
| | MDA | 12967.632[d] | 8 | 1620.954 | .677 | .711 | .049 |
| | AOPP | 7449.858[e] | 8 | 931.232 | .996 | .443 | .070 |
| Intercept | Folate | 4.163 | 1 | 4.163 | .300 | .585 | .003 |

| | | | | | | | |
|---|---|---|---|---|---|---|---|
| | GSH | 22.906 | 1 | 22.906 | 1.847 | .177 | .017 |
| | SH_THIOL | 68.014 | 1 | 68.014 | 5.286 | .023 | .047 |
| | MDA | 24992.792 | 1 | 24992.792 | 10.440 | .002 | .090 |
| | AOPP | 2966.404 | 1 | 2966.404 | 3.173 | .078 | .029 |
| Sex | Folate | 102.426 | 1 | 102.426 | 7.377 | .008 | .065 |
| | GSH | 15.298 | 1 | 15.298 | 1.233 | .269 | .012 |
| | SH_THIOL | 35.493 | 1 | 35.493 | 2.758 | .100 | .025 |
| | MDA | 2544.073 | 1 | 2544.073 | 1.063 | .305 | .010 |
| | AOPP | 2763.073 | 1 | 2763.073 | 2.956 | .088 | .027 |
| Age | Folate | 33.342 | 1 | 33.342 | 2.401 | .124 | .022 |
| | GSH | 4.233 | 1 | 4.233 | .341 | .560 | .003 |
| | SH_THIOL | .720 | 1 | .720 | .056 | .813 | .001 |
| | MDA | 5004.701 | 1 | 5004.701 | 2.090 | .151 | .019 |
| | AOPP | 11.977 | 1 | 11.977 | .013 | .910 | .000 |
| Lipid_drug | Folate | 27.675 | 1 | 27.675 | 1.993 | .161 | .018 |
| | GSH | 11.556 | 1 | 11.556 | .932 | .337 | .009 |
| | SH_THIOL | 94.834 | 1 | 94.834 | 7.371 | .008 | .065 |
| | MDA | 10.307 | 1 | 10.307 | .004 | .948 | .000 |
| | AOPP | 1666.822 | 1 | 1666.822 | 1.783 | .185 | .017 |
| HT_drug | Folate | 7.186 | 1 | 7.186 | .518 | .473 | .005 |
| | GSH | 4.273 | 1 | 4.273 | .345 | .558 | .003 |
| | SH_THIOL | 19.037 | 1 | 19.037 | 1.480 | .227 | .014 |
| | MDA | 131.543 | 1 | 131.543 | .055 | .815 | .001 |
| | AOPP | 2046.088 | 1 | 2046.088 | 2.189 | .142 | .020 |
| DM_drug | Folate | 29.513 | 1 | 29.513 | 2.126 | .148 | .020 |
| | GSH | 12.544 | 1 | 12.544 | 1.011 | .317 | .009 |
| | SH_THIOL | .581 | 1 | .581 | .045 | .832 | .000 |

| | | | | | | | |
|---|---|---|---|---|---|---|---|
| | MDA | 1030.454 | 1 | 1030.454 | .430 | .513 | .004 |
| | AOPP | 259.645 | 1 | 259.645 | .278 | .599 | .003 |
| vitamins | Folate | 8.120 | 1 | 8.120 | .585 | .446 | .005 |
| | GSH | .466 | 1 | .466 | .038 | .847 | .000 |
| | SH_THIOL | 54.671 | 1 | 54.671 | 4.249 | .042 | .039 |
| | MDA | 542.471 | 1 | 542.471 | .227 | .635 | .002 |
| | AOPP | 19.389 | 1 | 19.389 | .021 | .886 | .000 |
| Calcium | Folate | 11.099 | 1 | 11.099 | .799 | .373 | .007 |
| | GSH | 6.570 | 1 | 6.570 | .530 | .468 | .005 |
| | SH_THIOL | 94.616 | 1 | 94.616 | 7.354 | .008 | .065 |
| | MDA | 643.855 | 1 | 643.855 | .269 | .605 | .003 |
| | AOPP | 93.414 | 1 | 93.414 | .100 | .753 | .001 |
| fish_oil | Folate | 58.425 | 1 | 58.425 | 4.208 | .043 | .038 |
| | GSH | 1.989 | 1 | 1.989 | .160 | .690 | .002 |
| | SH_THIOL | 6.489 | 1 | 6.489 | .504 | .479 | .005 |
| | MDA | 43.836 | 1 | 43.836 | .018 | .893 | .000 |
| | AOPP | 29.723 | 1 | 29.723 | .032 | .859 | .000 |
| Error | Folate | 1471.762 | 106 | 13.885 | | | |
| | GSH | 1314.752 | 106 | 12.403 | | | |
| | SH_THIOL | 1363.874 | 106 | 12.867 | | | |
| | MDA | 253766.942 | 106 | 2394.028 | | | |
| | AOPP | 99089.133 | 106 | 934.803 | | | |
| Total | Folate | 20401.568 | 115 | | | | |
| | GSH | 4580.723 | 115 | | | | |
| | SH_THIOL | 17282.591 | 115 | | | | |
| | MDA | 2430801.202 | 115 | | | | |
| | AOPP | 1093201.332 | 115 | | | | |

| | | | |
|---|---|---|---|
| Corrected Total | Folate | 1728.295 | 114 |
| | GSH | 1371.115 | 114 |
| | SH_THIOL | 1676.274 | 114 |
| | MDA | 266734.573 | 114 |
| | AOPP | 106538.991 | 114 |

**Estimates**

| Dependent Variable | Lipid_Med | Mean | Std. Error | 95% Confidence Interval | |
|---|---|---|---|---|---|
| | | | | Lower Bound | Upper Bound |
| Folate | No | 13.348[a] | .552 | 12.254 | 14.441 |
| | Yes | 12.325[a] | .457 | 11.419 | 13.230 |
| GSH | No | 4.892[a] | .521 | 3.858 | 5.926 |
| | Yes | 5.553[a] | .432 | 4.698 | 6.409 |
| SH_THIOL | No | 12.769[a] | .531 | 11.716 | 13.822 |
| | Yes | 10.875[a] | .440 | 10.004 | 11.747 |
| MDA | No | 137.548[a] | 7.244 | 123.186 | 151.910 |
| | Yes | 136.923[a] | 5.995 | 125.038 | 148.809 |
| AOPP | No | 87.932[a] | 4.527 | 78.957 | 96.906 |
| | Yes | 95.871[a] | 3.746 | 88.444 | 103.299 |

No/Yes: use of antidiabetic drugs versus no drugs.

**ESF, Table 2.** Effects of medications and supplements on lipid variables.

**Multivariate Tests**[a]

| Effect | | Value | F | Hypothesis df | Error df | Sig. | Partial Eta Squared |
|---|---|---|---|---|---|---|---|
| Intercept | Pillai's Trace | .698 | 26.221[b] | 9.000 | 102.000 | .000 | .698 |
| | Wilks' Lambda | .302 | 26.221[b] | 9.000 | 102.000 | .000 | .698 |
| | Hotelling's Trace | 2.314 | 26.221[b] | 9.000 | 102.000 | .000 | .698 |
| | Roy's Largest Root | 2.314 | 26.221[b] | 9.000 | 102.000 | .000 | .698 |
| Sex | Pillai's Trace | .195 | 2.751[b] | 9.000 | 102.000 | .006 | .195 |
| | Wilks' Lambda | .805 | 2.751[b] | 9.000 | 102.000 | .006 | .195 |
| | Hotelling's Trace | .243 | 2.751[b] | 9.000 | 102.000 | .006 | .195 |
| | Roy's Largest Root | .243 | 2.751[b] | 9.000 | 102.000 | .006 | .195 |
| Age | Pillai's Trace | .066 | .806[b] | 9.000 | 102.000 | .612 | .066 |
| | Wilks' Lambda | .934 | .806[b] | 9.000 | 102.000 | .612 | .066 |
| | Hotelling's Trace | .071 | .806[b] | 9.000 | 102.000 | .612 | .066 |
| | Roy's Largest Root | .071 | .806[b] | 9.000 | 102.000 | .612 | .066 |
| Lipid_drug | Pillai's Trace | .143 | 1.890[b] | 9.000 | 102.000 | .062 | .143 |
| | Wilks' Lambda | .857 | 1.890[b] | 9.000 | 102.000 | .062 | .143 |
| | Hotelling's Trace | .167 | 1.890[b] | 9.000 | 102.000 | .062 | .143 |
| | Roy's Largest Root | .167 | 1.890[b] | 9.000 | 102.000 | .062 | .143 |
| HT_drug | Pillai's Trace | .084 | 1.039[b] | 9.000 | 102.000 | .415 | .084 |
| | Wilks' Lambda | .916 | 1.039[b] | 9.000 | 102.000 | .415 | .084 |
| | Hotelling's Trace | .092 | 1.039[b] | 9.000 | 102.000 | .415 | .084 |
| | Roy's Largest Root | .092 | 1.039[b] | 9.000 | 102.000 | .415 | .084 |
| DM_drug | Pillai's Trace | .132 | 1.724[b] | 9.000 | 102.000 | .093 | .132 |

| | | | | | | | |
|---|---|---|---|---|---|---|---|
| | Wilks' Lambda | .868 | 1.724[b] | 9.000 | 102.000 | .093 | .132 |
| | Hotelling's Trace | .152 | 1.724[b] | 9.000 | 102.000 | .093 | .132 |
| | Roy's Largest Root | .152 | 1.724[b] | 9.000 | 102.000 | .093 | .132 |
| vitamins | Pillai's Trace | .055 | .654[b] | 9.000 | 102.000 | .748 | .055 |
| | Wilks' Lambda | .945 | .654[b] | 9.000 | 102.000 | .748 | .055 |
| | Hotelling's Trace | .058 | .654[b] | 9.000 | 102.000 | .748 | .055 |
| | Roy's Largest Root | .058 | .654[b] | 9.000 | 102.000 | .748 | .055 |
| Calcium | Pillai's Trace | .079 | .979[b] | 9.000 | 102.000 | .462 | .079 |
| | Wilks' Lambda | .921 | .979[b] | 9.000 | 102.000 | .462 | .079 |
| | Hotelling's Trace | .086 | .979[b] | 9.000 | 102.000 | .462 | .079 |
| | Roy's Largest Root | .086 | .979[b] | 9.000 | 102.000 | .462 | .079 |
| fish_oil | Pillai's Trace | .033 | .381[b] | 9.000 | 102.000 | .942 | .033 |
| | Wilks' Lambda | .967 | .381[b] | 9.000 | 102.000 | .942 | .033 |
| | Hotelling's Trace | .034 | .381[b] | 9.000 | 102.000 | .942 | .033 |
| | Roy's Largest Root | .034 | .381[b] | 9.000 | 102.000 | .942 | .033 |

Lipid drugs: cholesterol lowering drugs, HT drugs: antihypertensive drugs, DM drugs: antidiabetic agents.

#### Tests of Between-Subjects Effects

| Source | Dependent Variable | Type III Sum of Squares | df | Mean Square | F | Sig. | Partial Eta Squared |
|---|---|---|---|---|---|---|---|
| Corrected Model | Cholesterol | 59415.532[a] | 8 | 7426.942 | 5.551 | .000 | .288 |
| | HDL cholesterol | 3713.932[b] | 8 | 464.242 | 2.366 | .022 | .147 |
| | LDL cholesterol | 37127.155[c] | 8 | 4640.894 | 4.164 | .000 | .232 |

| | | | | | | | |
|---|---|---|---|---|---|---|---|
| | Triglycerides | 46667.848[d] | 8 | 5833.481 | .894 | .524 | .061 |
| | ApoA | 13977.042[e] | 8 | 1747.130 | 2.945 | .005 | .176 |
| | ApoB | 9442.622[f] | 8 | 1180.328 | 2.867 | .006 | .173 |
| | APOBonAPOA | .498[g] | 8 | .062 | 1.961 | .058 | .125 |
| | FC | 2692.486[h] | 8 | 336.561 | 4.720 | .000 | .256 |
| | Zscore(LCAT) LCAT | 4.596[i] | 8 | .575 | .548 | .818 | .038 |
| | Zscore(RCTreal) | 12.980[j] | 8 | 1.623 | 1.670 | .114 | .108 |
| Intercept | Cholesterol | 39374.294 | 1 | 39374.294 | 29.429 | .000 | .211 |
| | HDL cholesterol | 3033.239 | 1 | 3033.239 | 15.459 | .000 | .123 |
| | LDL cholesterol | 16233.902 | 1 | 16233.902 | 14.566 | .000 | .117 |
| | Triglycerides | 11941.227 | 1 | 11941.227 | 1.829 | .179 | .016 |
| | ApoA | 15037.746 | 1 | 15037.746 | 25.347 | .000 | .187 |
| | ApoB | 5705.556 | 1 | 5705.556 | 13.861 | .000 | .112 |
| | APOBonAPOA | .172 | 1 | .172 | 5.399 | .022 | .047 |
| | FC | 1803.688 | 1 | 1803.688 | 25.295 | .000 | .187 |
| | Zscore(LCAT) LCAT | .437 | 1 | .437 | .416 | .520 | .004 |
| | Zscore(RCTreal) | .637 | 1 | .637 | .656 | .420 | .006 |
| Sex | Cholesterol | 12203.341 | 1 | 12203.341 | 9.121 | .003 | .077 |
| | HDL cholesterol | 1527.513 | 1 | 1527.513 | 7.785 | .006 | .066 |
| | LDL cholesterol | 2999.162 | 1 | 2999.162 | 2.691 | .104 | .024 |
| | Triglycerides | 335.577 | 1 | 335.577 | .051 | .821 | .000 |
| | ApoA | 6809.992 | 1 | 6809.992 | 11.479 | .001 | .094 |
| | ApoB | 779.411 | 1 | 779.411 | 1.893 | .172 | .017 |
| | APOBonAPOA | .031 | 1 | .031 | .975 | .326 | .009 |
| | FC | 627.136 | 1 | 627.136 | 8.795 | .004 | .074 |
| | Zscore(LCAT) LCAT | .453 | 1 | .453 | .432 | .512 | .004 |
| | Zscore(RCTreal) | 5.915 | 1 | 5.915 | 6.087 | .015 | .052 |

| | | | | | | | |
|---|---|---|---|---|---|---|---|
| Age | Cholesterol | 3261.867 | 1 | 3261.867 | 2.438 | .121 | .022 |
| | HDL cholesterol | 262.455 | 1 | 262.455 | 1.338 | .250 | .012 |
| | LDL cholesterol | 1237.369 | 1 | 1237.369 | 1.110 | .294 | .010 |
| | Triglycerides | 3055.937 | 1 | 3055.937 | .468 | .495 | .004 |
| | ApoA | 696.196 | 1 | 696.196 | 1.173 | .281 | .011 |
| | ApoB | 216.660 | 1 | 216.660 | .526 | .470 | .005 |
| | APOBonAPOA | .000 | 1 | .000 | .009 | .924 | .000 |
| | FC | 226.285 | 1 | 226.285 | 3.173 | .078 | .028 |
| | Zscore(LCAT) LCAT | .436 | 1 | .436 | .416 | .520 | .004 |
| | Zscore(RCTreal) | .475 | 1 | .475 | .489 | .486 | .004 |
| Lipid_drug | Cholesterol | 2876.591 | 1 | 2876.591 | 2.150 | .145 | .019 |
| | HDL cholesterol | 117.625 | 1 | 117.625 | .599 | .440 | .005 |
| | LDL cholesterol | 5575.824 | 1 | 5575.824 | 5.003 | .027 | .044 |
| | Triglycerides | 14886.050 | 1 | 14886.050 | 2.281 | .134 | .020 |
| | ApoA | 244.372 | 1 | 244.372 | .412 | .522 | .004 |
| | ApoB | 557.760 | 1 | 557.760 | 1.355 | .247 | .012 |
| | APOBonAPOA | .050 | 1 | .050 | 1.565 | .214 | .014 |
| | FC | 140.078 | 1 | 140.078 | 1.964 | .164 | .018 |
| | Zscore(LCAT) LCAT | 3.067E-5 | 1 | 3.067E-5 | .000 | .996 | .000 |
| | Zscore(RCTreal) | .004 | 1 | .004 | .004 | .949 | .000 |
| HT_drug | Cholesterol | 7578.131 | 1 | 7578.131 | 5.664 | .019 | .049 |
| | HDL cholesterol | 521.555 | 1 | 521.555 | 2.658 | .106 | .024 |
| | LDL cholesterol | 4195.735 | 1 | 4195.735 | 3.765 | .055 | .033 |
| | Triglycerides | 21.663 | 1 | 21.663 | .003 | .954 | .000 |
| | ApoA | 828.789 | 1 | 828.789 | 1.397 | .240 | .013 |
| | ApoB | 853.410 | 1 | 853.410 | 2.073 | .153 | .018 |
| | APOBonAPOA | .004 | 1 | .004 | .126 | .723 | .001 |

| | | | | | | | |
|---|---|---|---|---|---|---|---|
| | FC | 351.726 | 1 | 351.726 | 4.933 | .028 | .043 |
| | Zscore(LCAT) LCAT | .250 | 1 | .250 | .238 | .626 | .002 |
| | Zscore(RCTreal) | 1.041 | 1 | 1.041 | 1.071 | .303 | .010 |
| DM_drug | Cholesterol | 14076.205 | 1 | 14076.205 | 10.521 | .002 | .087 |
| | HDL cholesterol | 42.582 | 1 | 42.582 | .217 | .642 | .002 |
| | LDL cholesterol | 9203.380 | 1 | 9203.380 | 8.258 | .005 | .070 |
| | Triglycerides | 3426.451 | 1 | 3426.451 | .525 | .470 | .005 |
| | ApoA | 222.565 | 1 | 222.565 | .375 | .541 | .003 |
| | ApoB | 2977.946 | 1 | 2977.946 | 7.234 | .008 | .062 |
| | APOBonAPOA | .133 | 1 | .133 | 4.190 | .043 | .037 |
| | FC | 366.147 | 1 | 366.147 | 5.135 | .025 | .045 |
| | Zscore(LCAT) LCAT | .619 | 1 | .619 | .590 | .444 | .005 |
| | Zscore(RCTreal) | .715 | 1 | .715 | .735 | .393 | .007 |
| Vitamins | Cholesterol | 1141.546 | 1 | 1141.546 | .853 | .358 | .008 |
| | HDL cholesterol | 135.673 | 1 | 135.673 | .691 | .407 | .006 |
| | LDL cholesterol | 139.381 | 1 | 139.381 | .125 | .724 | .001 |
| | Triglycerides | 15018.092 | 1 | 15018.092 | 2.301 | .132 | .020 |
| | ApoA | 628.024 | 1 | 628.024 | 1.059 | .306 | .010 |
| | ApoB | 7.273 | 1 | 7.273 | .018 | .894 | .000 |
| | APOBonAPOA | .004 | 1 | .004 | .116 | .734 | .001 |
| | FC | 162.136 | 1 | 162.136 | 2.274 | .134 | .020 |
| | Zscore(LCAT) LCAT | 1.717 | 1 | 1.717 | 1.636 | .204 | .015 |
| | Zscore(RCTreal) | .047 | 1 | .047 | .048 | .827 | .000 |
| Calcium | Cholesterol | 5639.655 | 1 | 5639.655 | 4.215 | .042 | .037 |
| | HDL cholesterol | 128.950 | 1 | 128.950 | .657 | .419 | .006 |
| | LDL cholesterol | 3817.332 | 1 | 3817.332 | 3.425 | .067 | .030 |
| | Triglycerides | 13968.389 | 1 | 13968.389 | 2.140 | .146 | .019 |

| | | | | | | | |
|---|---|---|---|---|---|---|---|
| | ApoA | 226.915 | 1 | 226.915 | .382 | .538 | .003 |
| | ApoB | 1950.028 | 1 | 1950.028 | 4.737 | .032 | .041 |
| | APOBonAPOA | .128 | 1 | .128 | 4.014 | .048 | .035 |
| | FC | 337.310 | 1 | 337.310 | 4.731 | .032 | .041 |
| | Zscore(LCAT) LCAT | .531 | 1 | .531 | .506 | .479 | .005 |
| | Zscore(RCTreal) | .952 | 1 | .952 | .980 | .324 | .009 |
| fish_oil | Cholesterol | 604.847 | 1 | 604.847 | .452 | .503 | .004 |
| | HDL cholesterol | 12.638 | 1 | 12.638 | .064 | .800 | .001 |
| | LDL cholesterol | 2171.507 | 1 | 2171.507 | 1.948 | .166 | .017 |
| | Triglycerides | 2177.275 | 1 | 2177.275 | .334 | .565 | .003 |
| | ApoA | 200.646 | 1 | 200.646 | .338 | .562 | .003 |
| | ApoB | 476.342 | 1 | 476.342 | 1.157 | .284 | .010 |
| | APOBonAPOA | .042 | 1 | .042 | 1.316 | .254 | .012 |
| | FC | .143 | 1 | .143 | .002 | .964 | .000 |
| | Zscore(LCAT) LCAT | 1.133 | 1 | 1.133 | 1.080 | .301 | .010 |
| | Zscore(RCTreal) | .016 | 1 | .016 | .016 | .899 | .000 |
| Error | Cholesterol | 147175.880 | 110 | 1337.963 | | | |
| | HDL cholesterol | 21582.639 | 110 | 196.206 | | | |
| | LDL cholesterol | 122597.484 | 110 | 1114.523 | | | |
| | Triglycerides | 717998.018 | 110 | 6527.255 | | | |
| | ApoA | 65260.068 | 110 | 593.273 | | | |
| | ApoB | 45280.487 | 110 | 411.641 | | | |
| | APOBonAPOA | 3.495 | 110 | .032 | | | |
| | FC | 7843.553 | 110 | 71.305 | | | |
| | Zscore(LCAT) LCAT | 115.403 | 110 | 1.049 | | | |
| | Zscore(RCTreal) | 106.888 | 110 | .972 | | | |
| Total | Cholesterol | 5317221.000 | 119 | | | | |

| | | | |
|---|---|---|---|
| | HDL cholesterol | 451659.000 | 119 |
| | LDL cholesterol | 2084159.000 | 119 |
| | Triglycerides | 2010506.000 | 119 |
| | ApoA | 2927063.000 | 119 |
| | ApoB | 1003741.000 | 119 |
| | APOBonAPOA | 46.204 | 119 |
| | FC | 201171.262 | 119 |
| | Zscore(LCAT) LCAT | 119.999 | 119 |
| | Zscore(RCTreal) | 119.870 | 119 |
| Corrected Total | Cholesterol | 206591.412 | 118 |
| | HDL cholesterol | 25296.571 | 118 |
| | LDL cholesterol | 159724.639 | 118 |
| | Triglycerides | 764665.866 | 118 |
| | ApoA | 79237.109 | 118 |
| | ApoB | 54723.109 | 118 |
| | APOBonAPOA | 3.994 | 118 |
| | FC | 10536.039 | 118 |
| | Zscore(LCAT) LCAT | 119.999 | 118 |
| | Zscore(RCTreal) | 119.868 | 118 |

a. R Squared = .288 (Adjusted R Squared = .236)

b. R Squared = .147 (Adjusted R Squared = .085)

c. R Squared = .232 (Adjusted R Squared = .177)

d. R Squared = .061 (Adjusted R Squared = -.007)

e. R Squared = .176 (Adjusted R Squared = .116)

f. R Squared = .173 (Adjusted R Squared = .112)

g. R Squared = .125 (Adjusted R Squared = .061)

h. R Squared = .256 (Adjusted R Squared = .201)

i. R Squared = .038 (Adjusted R Squared = -.032)

#### Estimates

| Dependent Variable | Diabetes Med | Mean | Std. Error | 95% Confidence Interval Lower Bound | Upper Bound |
|---|---|---|---|---|---|
| Cholesterol | No | 211.632[a] | 3.617 | 204.465 | 218.800 |
| | Yes | 178.928[a] | 9.349 | 160.401 | 197.456 |
| HDL cholesterol | No | 60.099[a] | 1.385 | 57.354 | 62.844 |
| | Yes | 58.300[a] | 3.580 | 51.205 | 65.395 |
| LDL cholesterol | No | 130.724[a] | 3.301 | 124.182 | 137.265 |
| | Yes | 104.279[a] | 8.533 | 87.369 | 121.189 |
| Triglycerides | No | 104.489[a] | 7.989 | 88.657 | 120.320 |
| | Yes | 88.353[a] | 20.650 | 47.430 | 129.276 |
| ApoA | No | 155.250[a] | 2.408 | 150.478 | 160.023 |
| | Yes | 151.138[a] | 6.226 | 138.801 | 163.476 |
| ApoB | No | 91.325[a] | 2.006 | 87.349 | 95.301 |
| | Yes | 76.283[a] | 5.186 | 66.006 | 86.559 |
| APOBonAPOA | No | .609[a] | .018 | .574 | .644 |
| | Yes | .509[a] | .046 | .418 | .599 |
| FC | No | 40.734[a] | .835 | 39.079 | 42.389 |
| | Yes | 35.459[a] | 2.158 | 31.182 | 39.737 |
| LCAT | No | 80.682[a] | .266 | 80.154 | 81.209 |
| | Yes | 80.112[a] | .688 | 78.748 | 81.475 |
| Zscore(RCTreal) | No | .035[a] | .097 | -.158 | .228 |
| | Yes | -.198[a] | .252 | -.697 | .301 |

No/Yes: use of antidiabetic drugs versus no drugs.

**ESF, Figures 1.** Effects of the interaction between amnestic mild cognitive impairment (aMCI) X HADS-D groups (cut-off 8) on triglyceride levels [NH=normal controls (0) versus aMCI (1); the analysis is adjusted for age, sex and body mass index and performed using the residualized values after regression on the drug state variables].

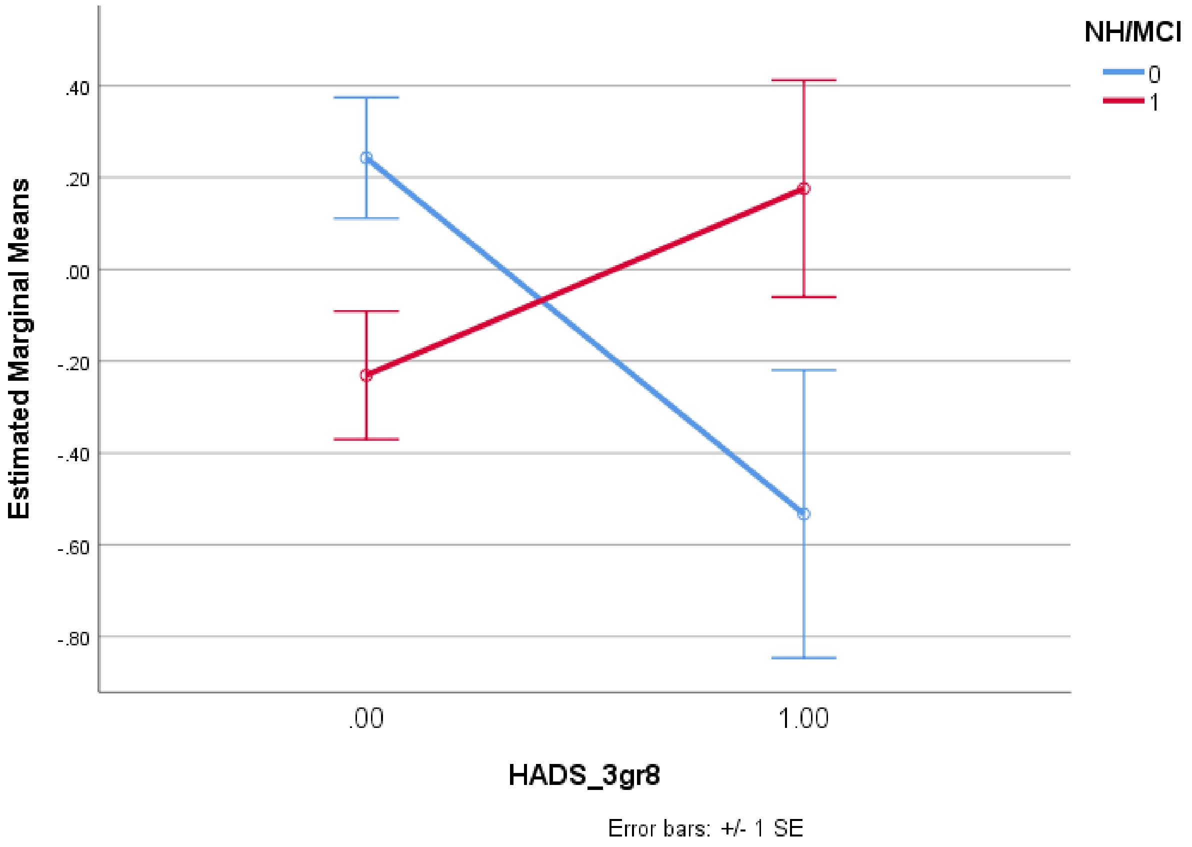

**ESF, Figures 2.** Effects of the interaction between amnestic mild cognitive impairment (aMCI) X HADS-D groups (cut off of 8) on high-density lipoprotein. [NH=normal controls (0) versus aMCI (1); the analysis is adjusted for age, sex and body mass index and performed using the residualized values after regression on the drug state variables].

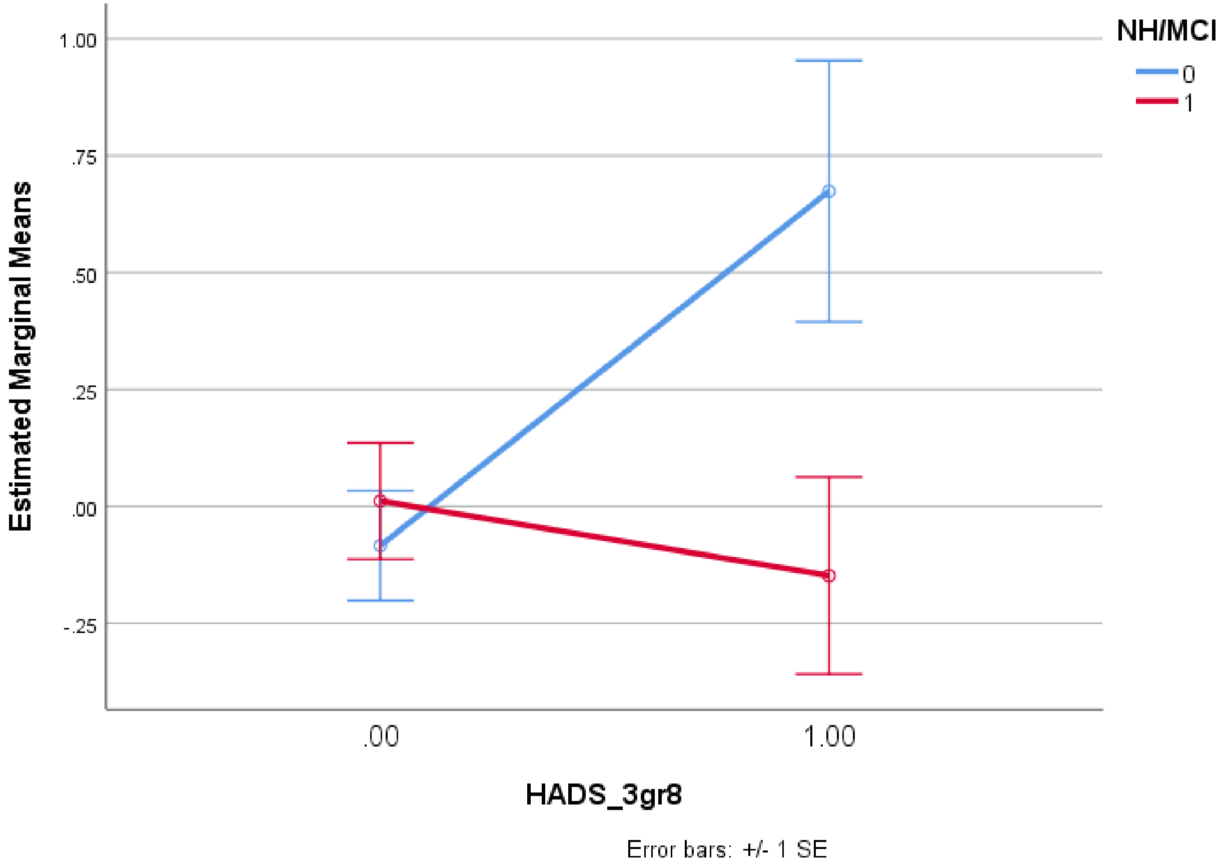

**ESF, Figures 3**. Effects of the interaction between amnestic mild cognitive impairment (aMCI) X HADS-D (cut off of 8) groups on apolipoprotein A [NH=normal controls (0) versus aMCI (1); the analysis is adjusted for age, sex and body mass index and performed using the residualized values after regression on the drug state variables].

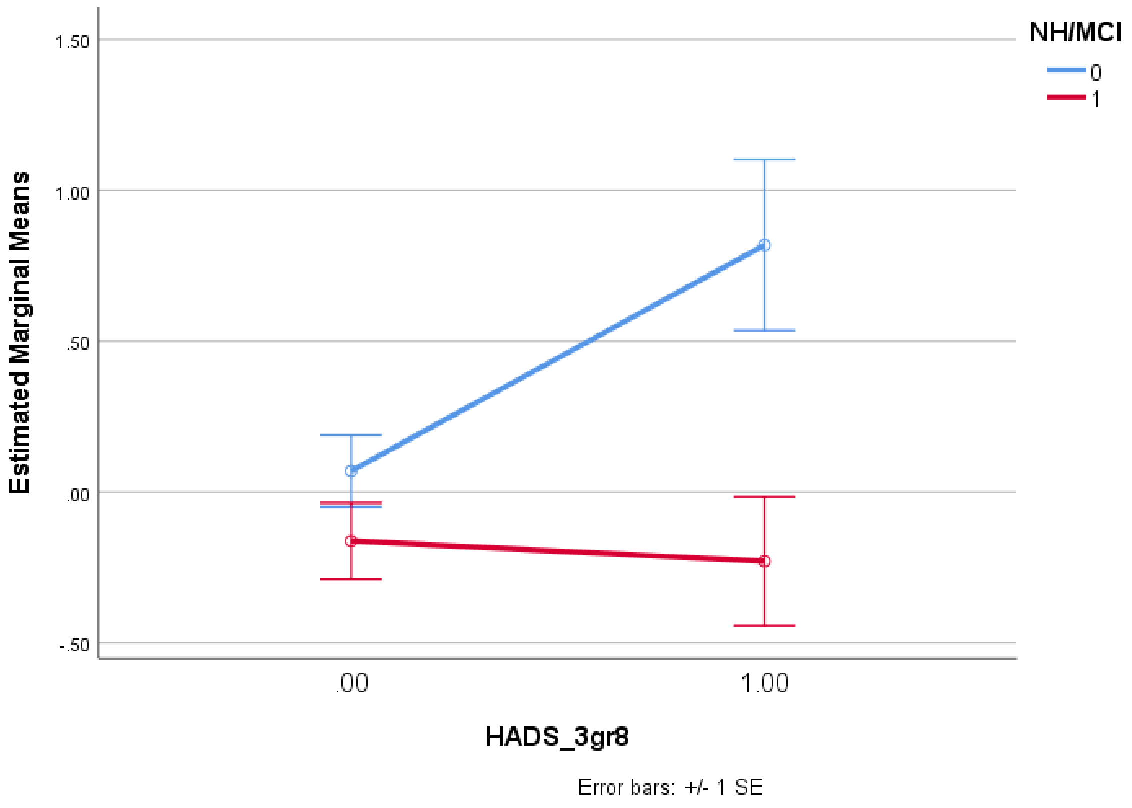

**ESF, Figures 4**. Effects of the interaction between amnestic mild cognitive impairment (aMCI) X HADS-D groups (cut off of 8) on apolipoprotein B. [NH=normal controls (0) versus aMCI (1); the analysis is adjusted for age, sex and body mass index and performed using the residualized values after regression on the drug state variables].

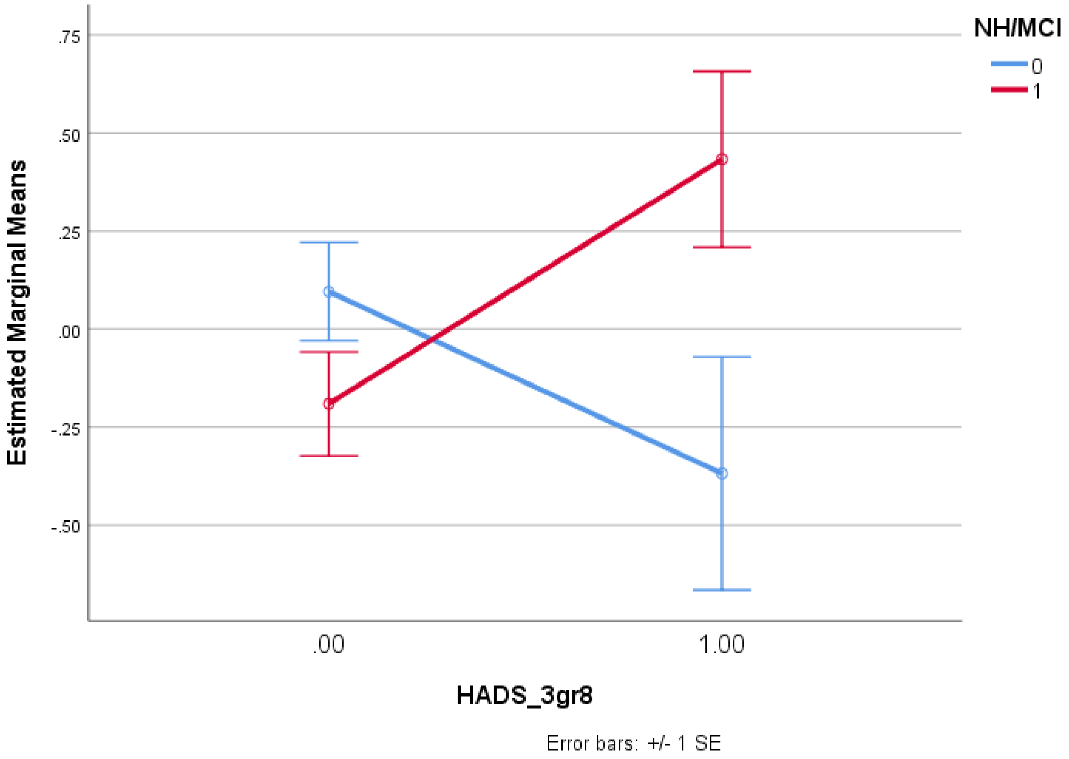

**ESF, Figures 6.** Effects of the interaction between amnestic mild cognitive impairment (aMCI) X HADS-D groups (cut off equals 8) on the apolipoprotein B (ApoB) / ApoB ratio [NH=normal controls (0) versus aMCI (1); the analysis is adjusted for age, sex and body mass index and performed using the residualized values after regression on the drug state variables].

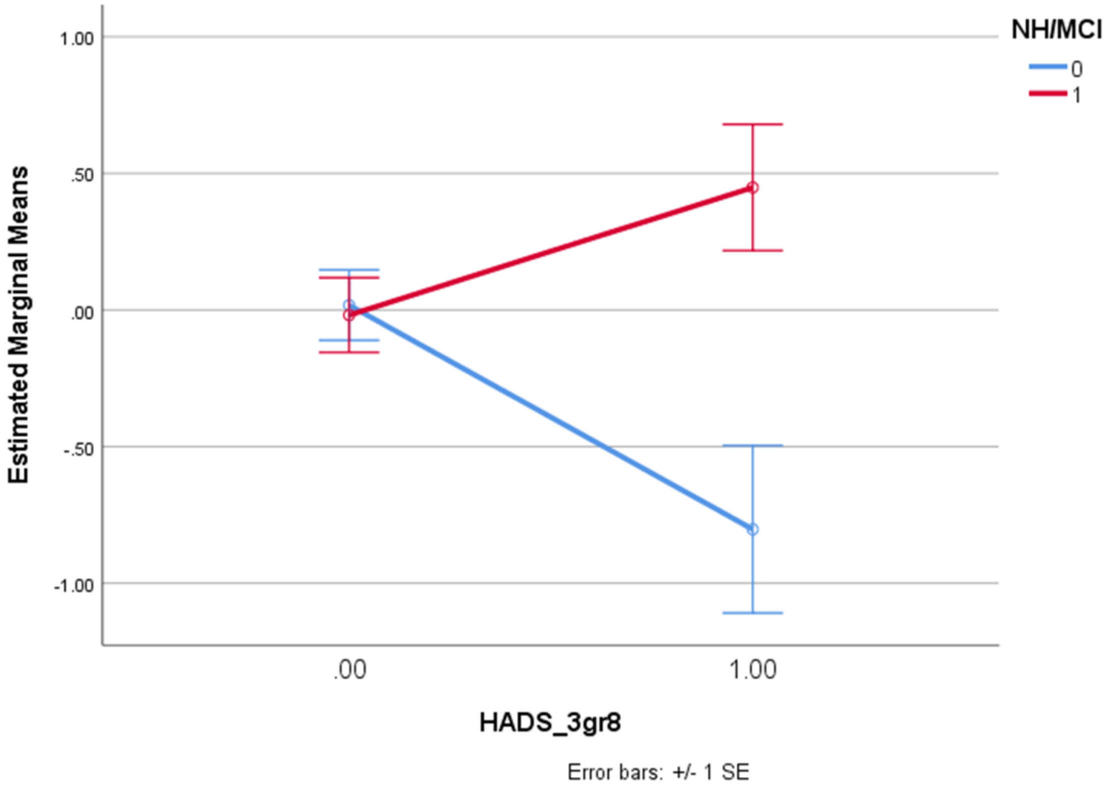

**ESF, Figures 6.** Effects of the interaction between amnestic mild cognitive impairment (aMCI) X HADS-D groups (cut off of 8) on the reverse cholesterol transport ratio [NH=normal controls (0) versus aMCI (1); the analysis is adjusted for age, sex and body mass index and performed using the residualized values after regression on the drug state variables].

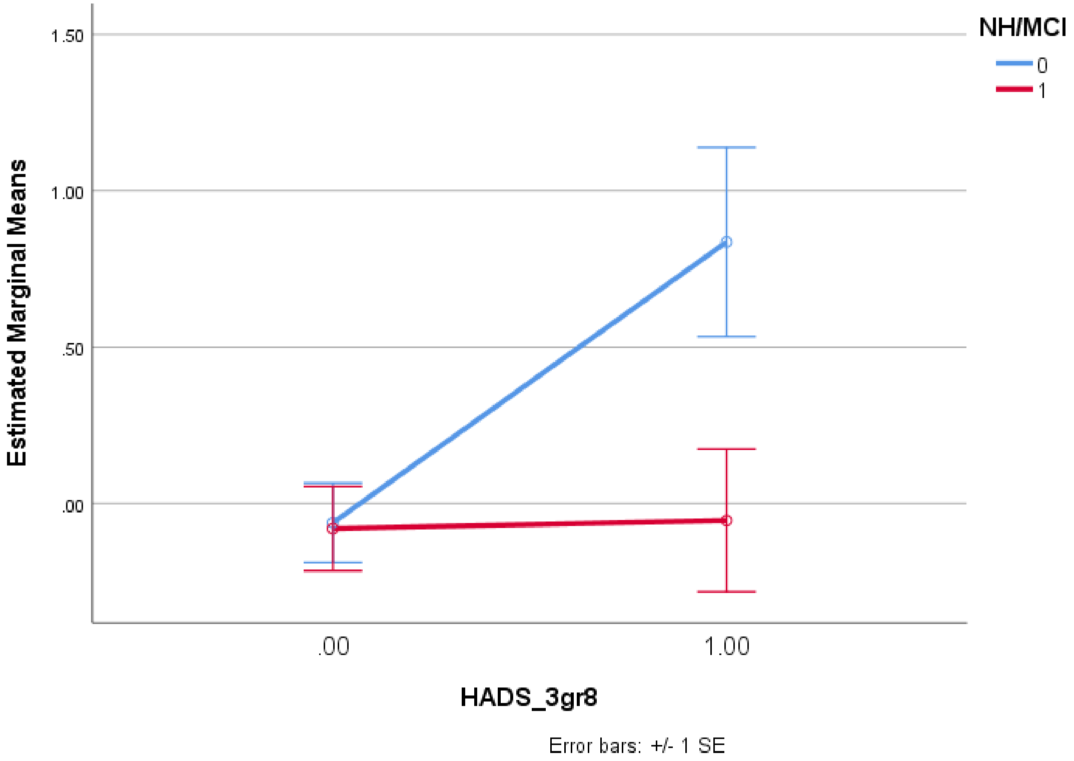

**ESF, Figures 7.** Effects of the interaction between amnestic mild cognitive impairment (aMCI) X HADS-D groups (cut off score of 8) on the Castelli risk index 1 [NH=normal controls (0) versus aMCI (1); the analysis is adjusted for age, sex and body mass index and performed using the residualized values after regression on the drug state variables].

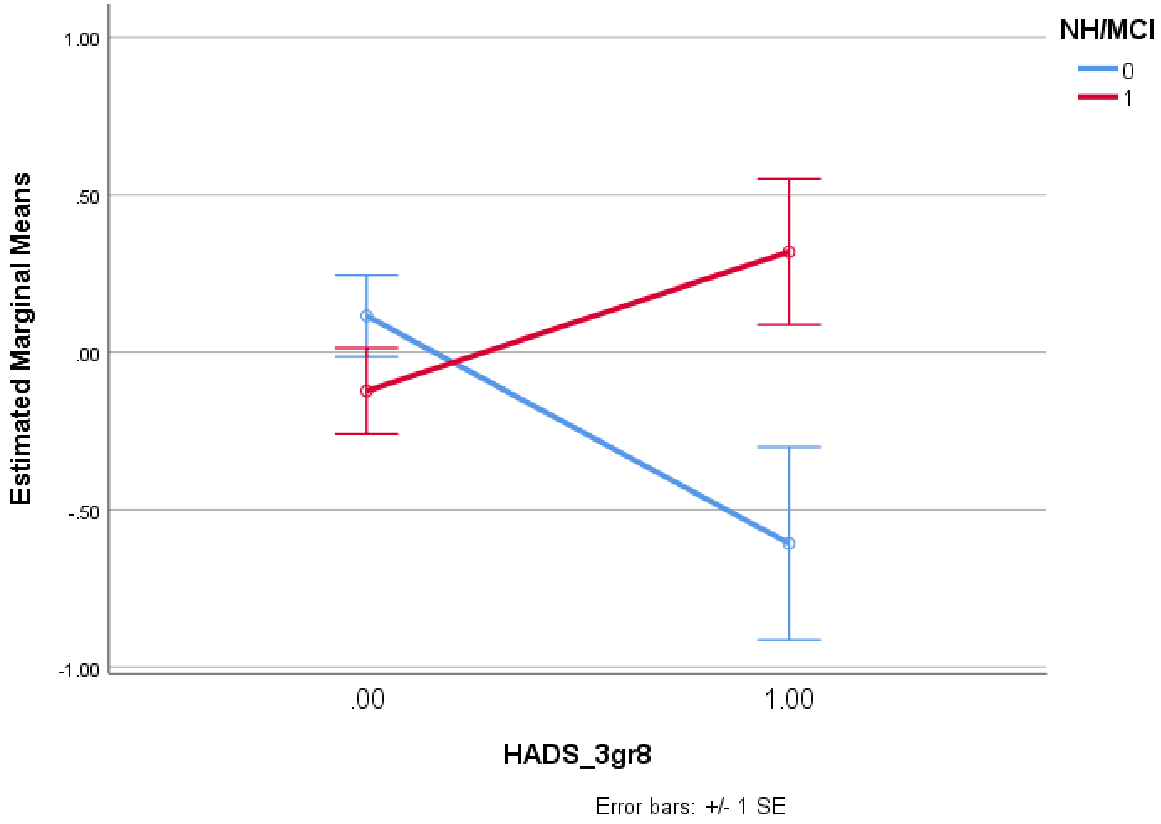
